# Empirical networks for localized COVID-19 interventions using WiFi infrastructure at university campuses

**DOI:** 10.1101/2021.03.16.21253662

**Authors:** Vedant Das Swain, Jiajia Xie, Maanit Madan, Sonia Sargolzaei, James Cai, Munmun De Choudhury, Gregory D. Abowd, Lauren N. Steimle, B. Aditya Prakash

**Affiliations:** College of Computing, Georgia Institute of Technology; H. Milton Stewart School of Industrial and Systems Engineering, Georgia Institute of Technology; Department of Computer Science, Brown University; College of Engineering, Northeastern University

**Keywords:** COVID-19, mobility, modeling, policy, non-pharmaceutical intervention, passive sensing, WiFi

## Abstract

Infectious diseases, like COVID-19, pose serious challenges to university campuses, which typically adopt closure as a non-pharmaceutical intervention to control spread and ensure a gradual return to normalcy. Intervention policies, such as remote instruction (RI) where large classes are offered online, reduce potential contact but also have broad side-effects on campus by hampering the local economy, students’ learning outcomes, and community wellbeing. In this paper, we demonstrate that university policymakers can mitigate these tradeoffs by leveraging anonymized data from their WiFi infrastructure to learn community mobility —- a methodology we refer to as *WiFi mobility models* (WiMob). This approach enables policymakers to explore more granular policies like localized closures (LC). WiMob can construct contact networks that capture behavior in various spaces, highlighting new potential transmission pathways and temporal variation in contact behavior. Additionally, WiMob enables us to design LC policies that close super-spreader locations on campus. By simulating disease spread with contact networks from WiMob, we find that LC maintains the same reduction in cumulative infections as RI while showing greater reduction in peak infections and internal transmission. Moreover, LC reduces campus burden by closing fewer locations, forcing fewer students into completely online schedules, and requiring no additional isolation. WiMob can empower universities to conceive and assess a variety of closure policies to prevent future outbreaks.

## Introduction

University campuses are often hotspots for infectious disease outbreaks and hence are targeted for interventions. In the wake of the Coronavirus Disease (COVID-19) [53], the U.S. witnessed more than half a million cases at universities [66], and colleges are still left with decisions for operations in Fall 2021 [39, 56]. Controlling the disease at universities can be pivotal to securing the surrounding environment [6]. To reduce on-campus infections and the likelihood of superspreading events, a recommended form of non-pharmaceutical intervention (NPI) is partial closure of the campus [22].

During COVID-19, advancement in teleconferencing technology equips universities to continue operations by adopting a form of campus closure that relies on remote instruction (RI) [49]. As a consequence, the campus community has fewer opportunities to visit spaces, such as classrooms, to congregate and risk transmission [1, 3]. One common approach campuses consider to design RI policies is to use enrollment data (En) to assume contact and therefore, offer large classes online while other classes remain in person [9, 72]. In fact, during COVID-19, 44% colleges and universities in the U.S., primarily offered instruction online [64]. However, these policies can still have broad, negative, and indiscriminate impact on the community by forcing students into completely remote course schedules. Such policies can have adverse effect on learning outcomes [18], where students can lose close to 7 months of education [2]. Additionally, RI can disincentivize students to stay on campus and thus, universities incur losses in auxiliary revenue (e.g., boarding, parking, dining, etc.) [25, 17], with universities standing to lose up to $50 million because of unused services [75]. Even the local population unaffiliated with the university takes sustains losses to business due to university closures [32, 71]. Furthermore, with socioeconomic disparities and heterogeneous household contexts, the demands of remote instruction can lead to added anxiety and stress among students [12, 74]. Relying on RI, university campuses struggle to balance community health with the demands of learning, economy, and broad wellbeing [58]. Instead, there is a need for a more versatile approach to design closure policies that empowers policymakers to accurately assess impact of closure interventions and model more data-driven targeted intervention strategies.

This paper showcases a new approach that universities can take to design closure policies by leveraging data from their existing WiFi infrastructure. Our methodology, *WiFi mobility models* (WiMob), involves constructing anonymized mobility networks of campus (Figure 1a), which helps determine extended periods of collocation — or “proximate contact” [30]— between individuals to describe contact networks on campus. Particularly, WiMob enables a more expressive toolkit for university policymakers that represents contact longitudinally and allows them to assess closure at the granularity of a room, suite, or hall. Thus, it lends itself to the design of targeted interventions that focus on localized closures (LC). We demonstrate the utility of WiMob with data collected over two years, of approximately 40, 000 anonymous occupants and visitors of the Georgia Institute of Technology (GT), a large urban campus in the U.S. — including about 16, 000 undergraduate students, 9, 000 graduate students, and 7, 600 staff members. In general, on comparing WiMob to En as an approach to model contact, we find that WiMob captures contact behavior at a community scale for a variety of campus spaces, describes temporal variations in contact, and provides a better estimate of local context by being aware of occupancy and the non-student population. Using WiMob also reveals that En overestimates the impact of RI on reducing contact on campus. Hence, we propose a less burdensome alternative to RI, by deriving more targeted LC policies based on WiMob (Figure 1) (indeed En is too coarse-grained for designing targeted LC policies).

**Figure 1:**
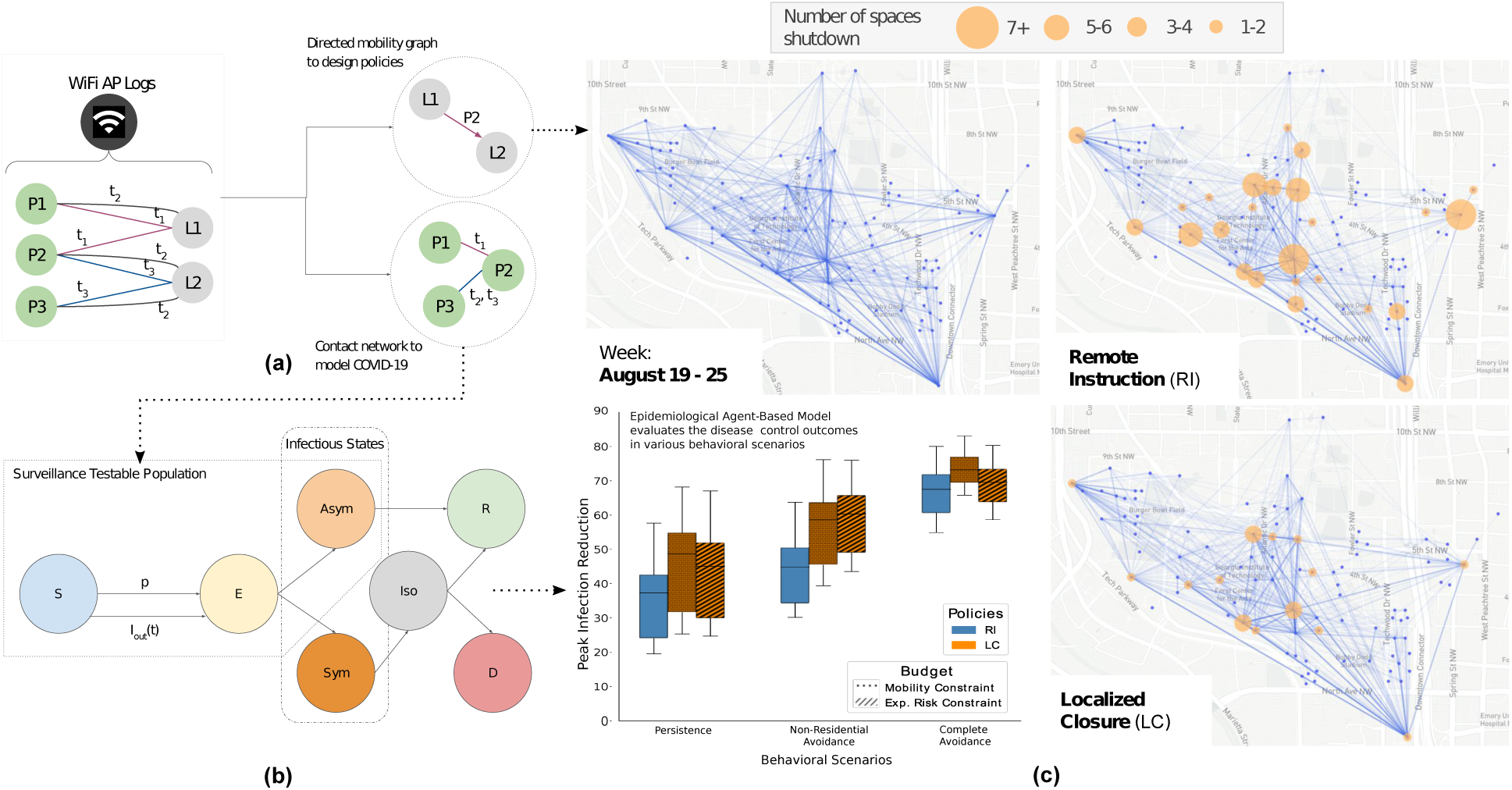
The WiFi mobility models (WiMob) methodology uses anonymized network logs to model campus mobility and target spaces for localized closures (LC) (*a*) WiFi network logs reflect timestamps when people’s devices associate with access points (APs) on campus. WiMob mines these logs to characterize mobility as a bipartite graph that describes people (e.g., *P* 1, *P* 2) visiting campus locations (e.g., *L*1, *L*2) during different times (e.g., *t*_1_, *t*_2_). Since people’s devices can proxy their presence, we estimate collocation (e.g., *P* 1 and *P* 2 were collocated at *L*1 at *t*_1_), and movement (*P* 2 dwelled at *L*1 and then at *L*2). (*b*) We use the collocation network construct a SEIR–based epidemiological ABM, calibrated to Fall 2020 incidence of COVID-19 (*c*) WiMob highlights mobility behavior to evaluate and inform policy. (*c*)–*top-left:* Mobility on campus between the top 100 most frequented locations on the GT campus in the Fall semester of 2019. Edges only connect points of significant dwelling and thus do not represent pedestrian routes. (*c*)–*top-right:* RI is a form of broad closure which affects a large number of students and locations.(*c*)–*bottom-right:* By contrast, we propose to use WiMob to parsimoniously identify a small set of spreader locations within buildings and design LC policies. (*c*)– *bottom-left:* We use our epidemiological ABM to evaluate these policies under different budgetary constraints and various behavioral scenarios (Persistence, Non-Residential Avoidance, Complete Avoidance). Our study shows that LC policies provide equal or better control on the disease spread, and yet minimize the burden on campus compared to RI.

We further exhibit that LC presents better disease control outcomes than RI by constructing and simulating an agent-based epidemiological model (ABM) over the people– people contact networks (Figure 1b) derived from the collocation identified with WiMob (Figure 1a). Our ABM was calibrated with GT on-campus COVID-19 cases from the Fall semester of 2020 [28] and infection rates from Fulton County [51]. To compare the effect of interventions, we construct a counterfactual semester — that is unaltered by other policy– induced behaviors of 2020 — by leveraging WiFi data from Fall 2019 to determine the contact structure of the simulation. We assess the effectiveness of closure NPIs (Figure 1c) by simulating COVID-19 under various behavioral scenarios. We find LC is comparable to RI in controlling total infections but more effective at reducing the peak infections and internal transmission. Additionally, LC targets fewer locations, forces fewer students into fully online schedules, and does not isolate any more people than RI – illustrating that WiMob can help universities devise highly-specific closure policies, like LC, which can contain disease spread and mitigate campus disruption in comparison to RI policies.

Our methodology also promises other advantages. Mobility generally has been used to dynamically model disease spread of influenza [59], rubella [73] and COVID-19 [3, 57] showing the effectiveness of mobility restrictions at a regional–, or city–level [79, 11, 7, 45, 34]. These studies typically rely on cell tower localization or aggregating GPS information from mobile phones [10]. Neither of these data sources is easy to access for university campuses. At the same time, studies to infer campus mobility networks have relied on accessing user devices with specialized data logging applications (e.g., contact tracing mobile apps) [13, 19, 61, 31], but these approaches are typically constrained for disease modeling because they require mass adoption to represent the entire community and continuous maintenance of software is needed to capture longitudinal behavior changes. In contrast, our work repurposes already existing managed WiFi networks to model mobility, which provides room level granularity for mobility [20, 70, 15, 67] and consequently indicates proximate contact [30]. Much like En, universities internally archive such data over a long term for other purposes and do not need to install any additional surveillance infrastructure to access it. Prior work has repurposed such data for campuses of size 10, 000 50, 000 in different locations including Singapore, the U.K., and the U.S [20, 76]. With the appropriate privacy considerations, a university can obtain such data at a low cost, continuously and unobtrusively. The possibility of pandemic still looms large in the future [37, 26]. As campuses prepare for the upcoming Fall semester and unforeseen contagious diseases of tomorrow, WiMob presents an attractive and practical method to inform better public health policies.

## Results

We present two sets of analyses in our work. The first set contrasts structural characteristics of contact networks described by WiMob with current practices that use enrollment data (En). In the next set, we used WiMob to build an epidemiological model (an agent-based model over the contact networks, referred to as ABM) and analyze the remote instruction (RI) and localized closure (LC) interventions in terms of their differences in dynamic disease-control outcomes and burdens to campus.

Note, throughout the paper we use the small-caps to denote different methodologies to model contact (WiMob and En) and sans-serif to denote different intervention strategies (RI and LC).

### WiMob provides local, holistic and dynamic structural insights for contact networks on campus

Studies on RI policies tend to assume that contact in universities is largely informed by En— transcripts showing which courses a student is registered for. En— can provide structural insights on density of connections and disease transmission paths to inform modeling disease simulations [29]. However, such static data can overestimate attendance and ignore overlap between courses (via instructors) and organic interactions outside classes (e.g., waiting areas, dining, parties, and extra-curricular activities). Therefore, using En can overemphasize the disease-mitigating structural changes to the network by RI interventions. By contrast, WiMob is more grounded in community behavior as it captures multiple scheduled and serendipitous contact situations dynamically over the semester. We compared the features of contact networks constructed with WiMob, against networks constructed with En using data from GT for Fall semester of 2019 (August 19 – December 14), prior to any COVID-19 reported cases in the U.S. En approximates contact based on students enrolling for classes that could potentially collocate them in the same room during lectures. WiMob infers contact when any two individuals actually collocate near the same WiFi access point [15, 67] for extended period (see explanation in SI WiFi Mobility). We found that WiMob rendered new insight into contact on campus that was invisible to the En methodology.

### WiMob characterizes temporal variation in proximity

Variation in contact over the semester would naturally impact the severity of disease spread. However, En describes a static network that does not capture such dynamics (Figure 2a). Instead, we found that WiMob shows contacts got sparser over the semester. Figure 2c presents a notable decline in contacts after the first two weeks, which coincides with multiple orientation seminars and the so-called “course shopping” period of Fall 2019. In fact, contact decreased considerably in classrooms, with a steeper slope possibly because of reduction in attendance. WiMob was able to reveal other observable changes, such as drop in contacts during exam period (week 15) and increase after fall recess (week 10). En rendered a highly connected static network, which can miscalculate the speed at which a disease spreads. By contrast, the longitudinal behavior represented by WiMob can help universities anticipate disease spread more accurately.

**Figure 2:**
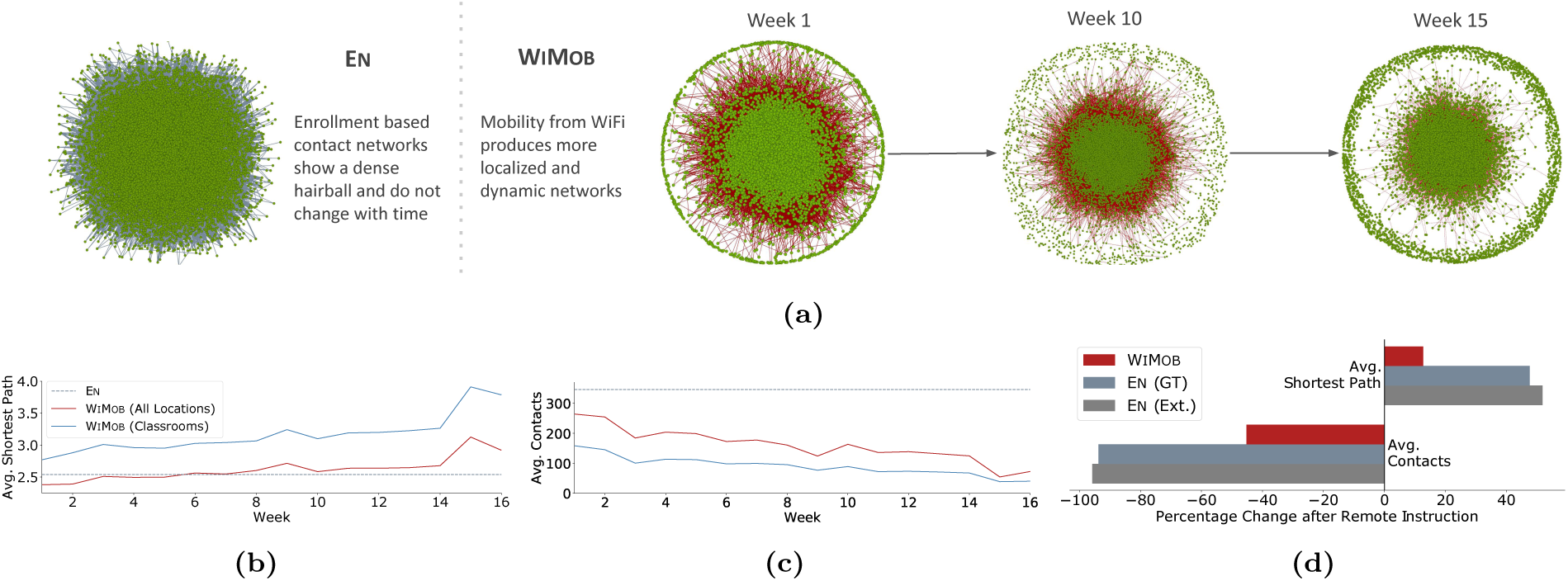
Results show difference in structural characteristics of contact networks from En (course enrollment) and WiMob (campus mobility). (*a*) In general, En overestimates connections (grey edges) between students (green nodes) and does not anticipate changes through the semester. En assumes 90% of students to be connected in a single component, but WiMob reveals (red edges) that on any given week only 69% are in the largest component (those not on campus are isolated and shown in the circumference). Moreover, WiMob reveals that density of connections changes over the semester. (*b*) En depicts campus contacts to be connected closely into a “small world”. WiMob shows that contacts evolve over time. As mobility captures interactions outside classrooms we observe that for the first 6 weeks the shortest transmission path between people is shorter than what is reported by En. (*c*) Enrolling into a course does not necessitate physically collocating with the class for extended periods (students can also choose to be entirely absent). WiMob reflects this behavior and highlights a decline in average contacts over time. (*d*) These structural differences can help policymakers anticipate the effect of closure policies by describing how it fragments the underlying contact network. En shows that remote instruction leads to a 94% reduction in contacts and 50% increase in transmission path length (similar to numbers reported in prior work [72], shown as En (Ext.)). However, the estimate is significantly lower when measured using WiMob. As a result, WiMob emphasizes the limits of remote instruction policies and in turn motivates new policies that can be designed and evaluated with actual on-campus behavior.

### En overestimates contact-based risk

Campuses can assess risk of an outbreak by characterizing the number of individuals that would be at risk of infection through contact. In our study, En indicated 99% of the individuals on campus were clustered in a single component — if any of them would have been infected in Fall 2019, the entire component would be at risk. From the lens of En a virus can exhaust an entire population with infection very early. However, WiMob showed that only 69% of the population was connected in a single component (Table S2). This difference is because WiMob can distinguish how many individuals are active on campus. Therefore, WiMob provides a pragmatic estimate of risk by grounding it in local occupancy and helps campuses budget for resources better.

### WiMob reveals different paths for disease transmission

Reports suggest that a key contributor to cases in the pandemic is actually clustering of individuals in non-academic spaces [49]. However, En does not depict a holistic view of campus contact. It is limited to classrooms and, therefore, fixates on contacts in lectures, while ignoring other spaces. In fact, WiMob showed that in the first 6 weeks of Fall 2019, the shortest path among individuals was smaller than that approximated by En (Figure 2b). With WiMob, we observed new paths in the contact network from situations outside classes. On a given week, WiMob showed the average shortest path with contact is 3.26(±0.5) when only considering lectures, whereas capturing all contexts reduced the average shortest path to 2.67(±0.28). Characterizing shorter pathways is crucial for policymakers as closure policies by design aim to disconnect these pathways.

### En overemphasizes the impact of remote instruction

Prior work uses En to posit that RI reduces contact and in turn significantly fragments the network for disease spread in universities [72, 9]. To compare policy effectiveness with WiMob, we operationalize RI in our study:

#### Remote Instruction (RI)

The status quo for data-driven policies offers strictly online instruction for large class enrollment, while continuing the other classes in person. When using En to model contacts, we implemented RI by removing connections between students who were only in contact through courses of size ≥ 30. When using WiMob to model contacts, we removed connections between students if they were only connected because of collocations during scheduled lectures of such courses.

We evaluated the effectiveness of such a policy if it were applied in Fall 2019, with both WiMob and En. Figure 2d shows that RI with En reduced contact by 94% and increases shortest path by 50%. However, the same intervention with WiMob showed a relatively milder impact (contact reduction 45%; shortest path increase 11%). This reinforces that contact outside courses are significant and remain unaffected by enrollment-oriented policies like RI. WiMob provides a more encompassing view of the structural effects to a network and motivates design of more impactful closure policies.

### Epidemiological model built with WiMob shows that LC yields better infection reduction outcomes with lower burden

As outlined above, En does not comprehensively capture the contact on campus. By contrast, contact networks built with WiMob demonstrate new structural insights, which are critical to describe disease spread. A campus is composed of many different spaces, and En does not have the flexibility to design closure of such spaces or assess its impact. These drawbacks naturally motivate a new approach to design interventions. Since WiMob mitigates the limitations of En, we leveraged it to demonstrate the effectiveness of localized closure (LC) policies.

We used WiMob to define the contact structure of each day and simulate COVID-19 with an agent-based model. Our ABM was overlayed by a modified SEIR compartmental model for COVID-19 for each agent. GT also had implemented a robust surveillance program on campus. Hence we calibrated the ABM on the positivity rate for COVID-19 for GT [28] in the first 5 weeks of Fall 2020 also incorporating external seeding from the surrounding Fulton County, GA [51]. We validated our model by predicting future trends for the rest of Fall 2020. For robustness, we performed additional calibrations by varying time windows and university context (details in SI Sensitivity Analyses). We studied interventions by applying the ABM over the contact networks produced by WiMob with data from Fall 2019 — a counterfactual to Fall 2020 if no closure had occurred (see SI Simulation Model for further details).

### WiMob can model RI and LC interventions with various configurations

Prior works show a few locations are responsible for majority spread [11] and restricting movement between them leads to greater control [38]. In addition to RI, we modeled LC, which we formalize as follows:

#### Localized Closure (LC)

We identified rooms–level spaces that are highly central location nodes in the network. We removed contacts between people who are only connected because of collocating at these locations. While, we employed various centrality algorithms to identify such locations, for the results discussed in this section we use *PageRank* [54]). Details in SI Identifying Locations for Closure.

We found that, if COVID-19 spread through Fall 2019 (a regular semester), the cases rose after 7 days (Figure 3a). Therefore, we applied both RI and LC interventions after the first week.

**Figure 3:**
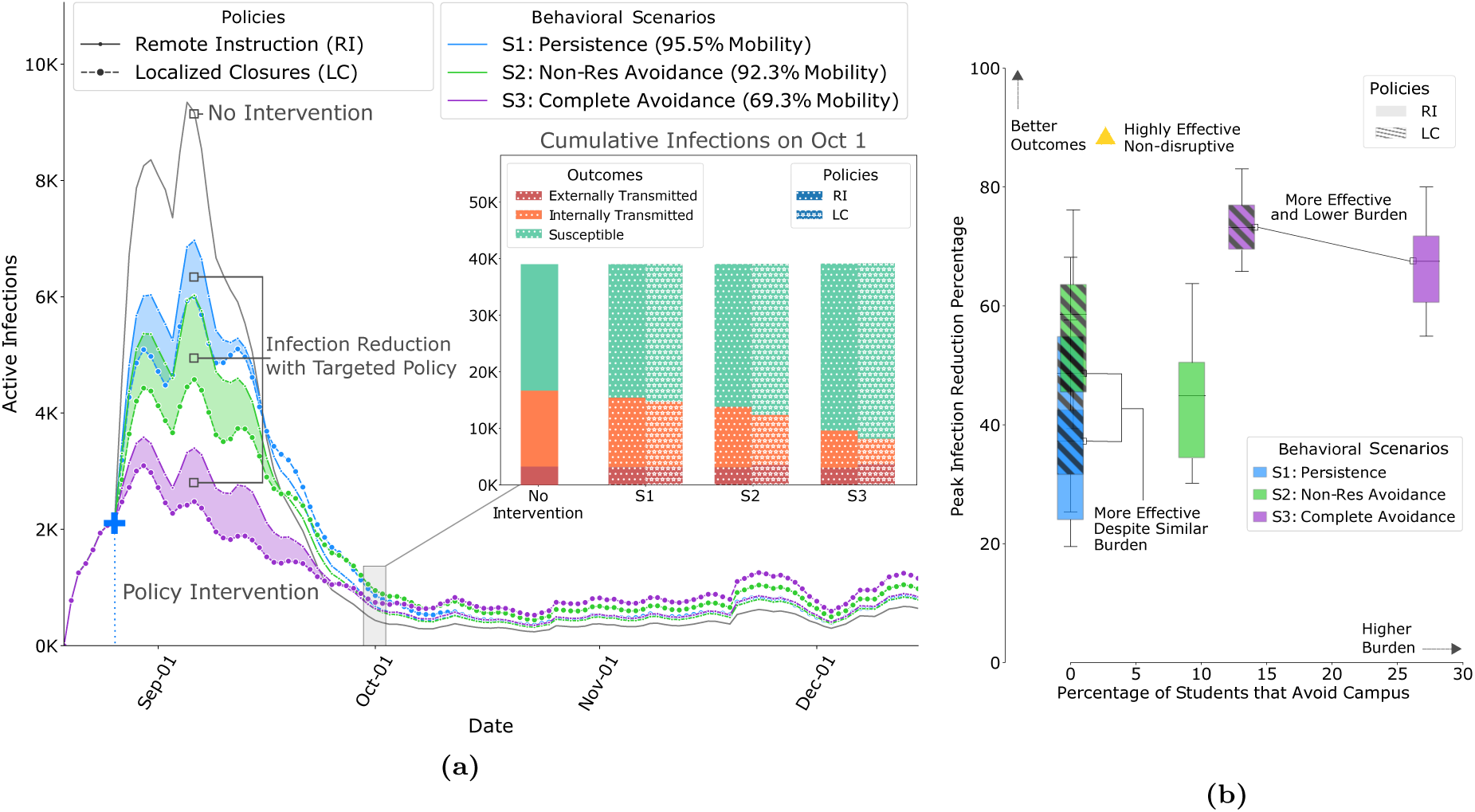
Results of policy interventions with our calibrated ABM on contact networks from Fall 2019, derived from WiMob (*a*) This graph compares the mean active infections between LC and RI. LC show improved outcomes (shaded regions) even when constrained to the same restrictions of RI policies. (*a*)–*inset:* After the first wave, even though LC shows slightly higher active infections, the cumulative infections are still lower, especially those that are a result of internal transmission on campus. Figure S11— Figure S18 show changes in cumulative infections under different policies, including 2.5*^th^* and 97.5*^th^* percentile intervals. (*b*) Outcomes of policies within the same behavioral scenario are shown with boxes of the same color (RI policies are solid, LC policies are hatched) and box heights represent the 2.5*^th^* and 97.5*^th^* percentile. In *S1*, even though LC and RI are equally burdensome in terms of students avoiding campus, LC shows improved outcome on peak reductions. In fact, for the other scenarios, LC shows better outcomes than RI, without forcing as many students into online schedules, and, therefore, being even less burdensome with greater impact. Figure S7— Figure S10 show comparison of all policy outcomes with different budgets.

To make the comparisons between the closure policies, we established fixed budgets to design LC based on the resource utilization on RI. We considered 2 kinds of budgets, (i) mobility reduction — to depict space use on campus, and (ii) risk of exposure — to reflect testing capacity. Also note, response to closure policies can lead to unpredictable side-effects in campus behavior, particularly when a student’s schedule is entirely online. Therefore, we design policies within three behavioral scenarios (each with a varying budget):

#### S1: Persistence

Irrespective of the locations closed or classes restricted, individuals continue their other visiting behaviors.

#### S2: Non-Residential Avoidance

Non-residential students stop all visits to campus if they enrolled in at least 3 courses and the policy forces their entire academic schedule online.

#### S3: Complete Avoidance

Same as S2, but even residential students avoid campus based on their schedule.

Similar to other works that model closure [72, 5], we assume that when a location is shutdown, the individuals who ought to have visited that location isolated during the time.

To devise interventions,WiMob estimated how RI uses the budget and then designed LC to match this budget under every behavioral scenario Table 1 describes how the budget for each policy varies. Additional details are present in SI Modeling Policy and Scenarios.

**Table 1:**
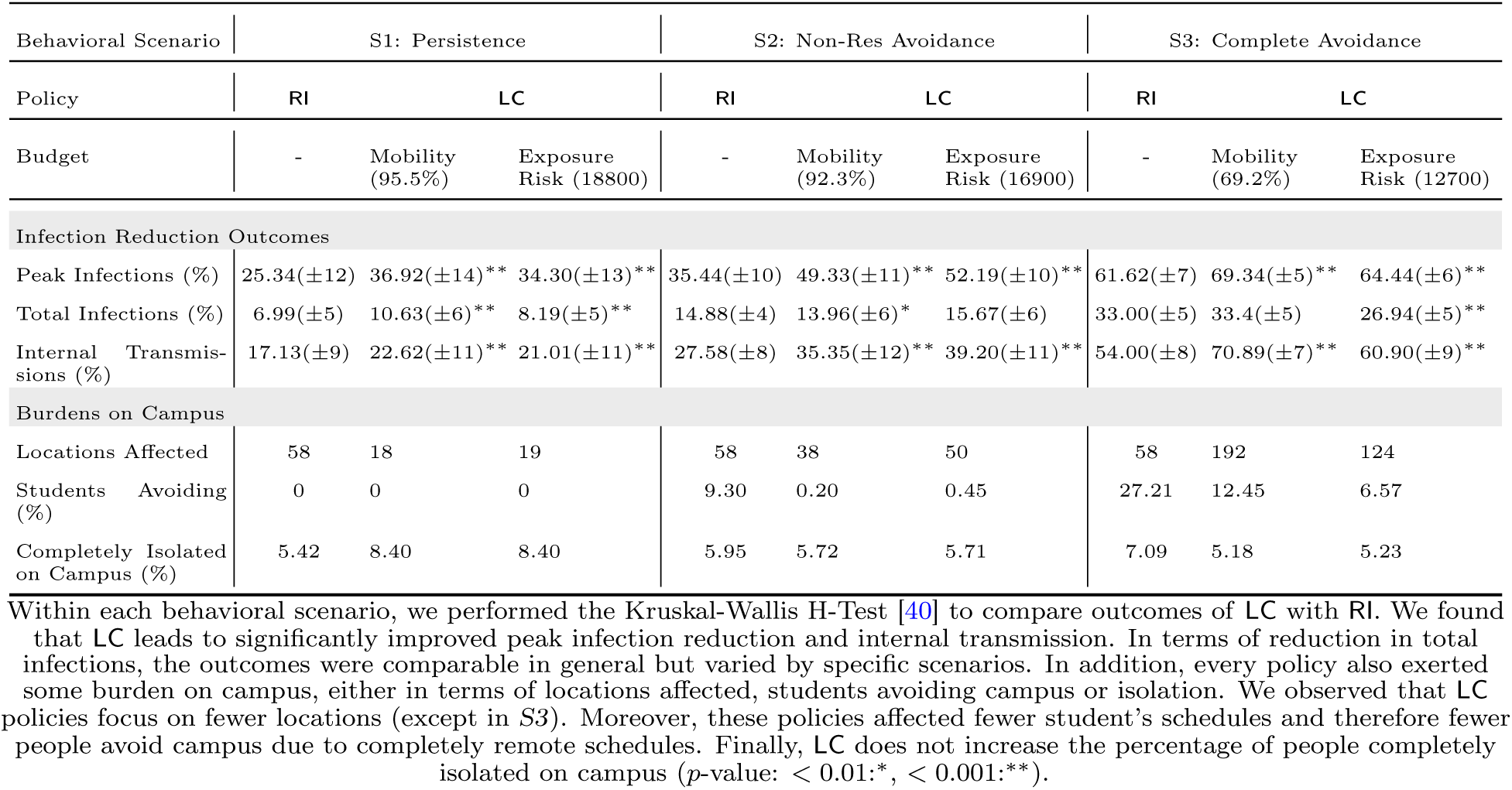
Comparison of policies in terms of controlling the disease and impacts on campus in Fall 2019.

We present differences between LC and RI based on three infection reduction outcomes; peak infections (maximum active cases on a given day), internal transmission (exposure from infected individuals on campus), and total infections (cumulative cases at the end of the semester). Additionally, we measured the burden of policy interventions with the number of locations closed — requires resources to monitor and maintain super-spreader locations, the percentage of students that avoid campus — disruption to learning outcomes [18, 12], and the percentage of individuals completely isolated — worsens mental wellbeing [60].

### LC cause greater reduction in peak infections, while affecting fewer locations

Controlling peak infections relaxes the burden on a university to support positive cases for any given day, and allows resources to be distributed over time. In all behavioral scenarios of our simulation of Fall 2019, we observed that the peak reduction was significantly better in LC (Figure 3) than RI. While RI impacted 58 different locations (classrooms and lecture halls), in *S1* and *S2*, LC achieved better outcomes by closing fewer locations. For example, in *S2*, RI achieved a 28.9% peak reduction, but LC showed reductions of 49.3% (mobility budget) and 48.1% (exposure risk budget). This was attained by closing 38 or 50 locations respectively. Therefore, with such policies, policymakers need to restrict fewer locations to remarkably minimize the pressure of active infections on campus (e.g., diagnoses, treatment, quarantining).

### LC lead to comparable reduction in total infections, while keeping more students on campus

Universities want to minimize the number of infected cases while ensuring majority of the population remains active on campus to continue successful operation. In Scenario *S1*, the total number of infections reduced by both LC was more than the reduction shown by RI. were similar. For other behavioral scenarios the total infection reduction between policies was similar ((Table S2). In contrast, the impact the policies had on the student schedules was remarkably different. RI forced multiple students to adapt to fully online schedules. In Scenario *S2*, 9% of students did not visit campus and in *S3*, 27% of students did not visit campus. On the other hand, in LC, the number of students expected to avoid campus could be as low as 0 and never exceeded 12%. Besides sustaining economic loss to the campus, remote instruction can increase anxiety among students and hinder learning outcomes [12, 74]. Compared to RI, LC offers policymakers a way to defend against turnover in the student population, without compromising overall control of disease spread (Table 1). Limiting the number of students that avoid campus helps preserve on-campus businesses [32, 71] and minimally disrupts the student wellbeing.

### LC cause greater reduction in internal transmission without causing further isolation on campus

Universities are responsible for limiting spread on campus, but they must also ensure that aggressive policies do not worsen mental wellbeing of the community. In terms of internal transmission the reduction is significantly larger with LC (Table 1). However, when LC restricted the infections early in Fall 2019, it left more individuals susceptible to external transmission. College student behavior outside campus on weekends and breaks is known to impact local transmission [16]. When policymakers consider LC they should also consider policies on re-entry or required testing based on off-campus activities. In terms of isolating individuals on campus, it’s notable that LC and RI were similar in *S2*. Interestingly, in *S3*, where LC closed more than 100 locations, the percentage of isolated individuals per week was less than that of RI. This finding implies that LC can keep individuals on campus without forcing them into complete isolation. Here “isolation” refers to no form of proximate contact with any individual on campus — extreme social distancing where individuals are not even collocated in the same suite or hall. While social distancing is a recommended countermeasure for COVID-19 [1], complete isolation can have adverse effects on psychological wellbeing [60, 41, 55]. LC can help alleviate concerns of closure interventions that increase loneliness and limit social connectedness [41].

### LC identifies a wider variety of auxiliary spaces

By using WiMob to design LC we were able to identify locations for closure at the granularity level of rooms, including unbound spaces such as lobbies and work areas. As policy design budgets changed with every behavioral scenario we found that LC identified different types of locations for closure. First, in *S1*, we found that most locations that LC targeted are a subset of the auditoriums–like rooms where large classes would take place in Fall 2019. Note, LC needs to restrict only a few such spaces to utilize the same budget as RI. This is because, under *S1*, RI policies only altered visits to lectures, while these spaces are used for other purposes during other times (e.g., club activities and seminars). We also noted that LC targeted ‘high traffic’ locations like conference center lobbies which are typically used as waiting areas or for networking events. Next, in Scenario *S2*, we saw that in addition to spaces mentioned earlier, interestingly LC further restricted the use of smaller rooms (occupancy 13 − 35) which would not be affected by RI (as only classes of size ≥ 30 are offered online). LC also targeted areas in the recreation center (which includes locker rooms and indoor courts for 4 − 20 people). This insight indicates that our methodology WiMob accounts for a diverse set of student activities. Moreover, we also found a selection of spaces that would not be frequented by the undergraduate population, such as lab areas and facility buildings like the police station. Lastly, in Scenario *S3*, LC targeted closure of activity in far more spaces than RI. However, the better outcomes can be attributed to the fact that LC diversified the potential restriction areas. LC restricted heavily used small study rooms or breakout rooms (for 1 − 6 people). Furthermore, it restricts use of spaces where multiple small groups of people can organically assemble, such as cafes, dining halls, and reading areas. We also observed that LC restricted activity in about 10 Greek Houses but does not target other housing areas — demonstrating its ability to restrict social behavior that could amplify disease spread. Figure S19 shows the diversity in locations for various LC policies.

### Sensitivity and robustness analyses

The results above use an ABM calibrated on the positivity rate of the first 5 weeks of Fall 2020. This rate can be influenced by many latent factors (e.g., mask-wearing, hand washing, distancing, and compliance). To study any effect of these variations, we also calibrated on different time windows throughout the semester. We calibrate on weeks 5 9 and 10 14 in Fall 2020, and validate on the remaining semester. In both cases, compared to RI, we found that LC still exhibits better reduction in peak infections (up to 90%) and internal transmission (up to 77%). In the original calibration, LC maintained the same level of total infections as RI, but with the new periods we found total infections were substantially less than RI (Table S8 and Table S9). Another important variable for positivity is the wider context of the campus e.g. urban/rural, the surrounding county, city, etc. To investigate this, we also calibrated our ABM on the positivity rate of different universities in the US in Fall 2020 (along with information from their county to seed external cases). Consider this as a hypothetical where the mobility of the GT community remains the same but disease outcomes resemble a different campus. We calibrated on data from University of Illinois at Urbana-Champaign and University of California, Berkeley. We found no remarkable differences from our findings with GT (Table S10 and Table S11).

## Discussion

Non-pharmaceutical interventions (NPI) are the first line of defense for universities to respond to contagious diseases like COVID-19 [21, 47] and are also crucial to control infections and continue operations until recovery. On a campus, a common form of NPI is closure [33]. Universities consider enrollment data (En) to design remote instruction (RI) for closure to support continued operations safely [72]. However, En can misconstrue contact on campus, and RI policies can have broad impacts despite their effects on curbing the disease spread. This paper demonstrates that repurposing logs from a managed WiFi network (WiMob) can help design effective localized closure policies (LC). We show that WiMob uncovers rich contact dynamics and provides policymakers multiple dimensions to design policies like LC. We simulate COVID-19 with an ABM that harnesses WiMob to compare RI and LC. As universities plan for Fall 2021, our results present evidence that LC designed with WiMob can lead to improved infection reduction outcomes, while simultaneously relaxing burdens on the campus caused by coarse-grained broad RI policies.

### Generalizability for Other Contexts

In practice LC policies should be deployed in conjunction with the other tools as well like testing, tracing, and quarantining. WiMob can complement disease-specific knowledge to identify closure spaces. For example, small indoor spaces with poor ventilation increase the risk of infection for COVID-19 [63], while other algorithm-identified locations for closure might not require closure because users of a space are compliant with mask-wearing and testing. Further, as a pandemic progresses and public health guidance develops [62], with WiMob, campuses can regulate the restriction of LC policies and anticipate the path to ‘normal’ operations [39, 56]. Moreover, WiMob captures various spillover effects that cannot be captured in methods like En. For instance, with WiMob we observe that the mobility in Fall 2020 was 39% of that in Fall 2019 because the on-ground policies lead to certain staff working remotely as well. With additional information, WiMob enables policymakers to model such scenarios and design alternatives like LC with new budgets. Policymakers can use WiMob as a versatile tool to explore dynamic intervention strategies as well. Prior work shows that staggering policy restrictions could have variable impact on campus [78]. Accordingly, WiMob could be used to build an adaptive version of LC that updates at different points in the semester based on expected mobility changes. Additionally, depending on campus priorities and resource limitations, different campuses can use this same data to model policies differently. The effectiveness of reopening policies is expected to be sensitive to a campus’ specific context that includes physical infrastructure, overarching guidelines, and human compliance [6]. For certain campuses policies might not need to be constrained by exposure risk as testing might be frequent, ubiquitous, and voluminous. Other campuses could have limits on quarantining capacity. Policymakers might even consider the cost trade-offs by actually forecasting actual financial losses incurred by reduction in mobility [7], or valuate loss of services based on community needs [65]. We elaborate on these considerations in the SI Implications for Policy Design.

### Operational Considerations

Beyond assessing cost-benefits, universities need to devise practical methods of obtaining, storing, and processing mobility of the community as WiMob. University can access logs from the managed network internally as it is passively collected. Moreover, it does not require any new form of surveillance sensing but universities must revise terms of use and stay sensitive to community perspectives. Despite population mobility being valuable for many applications [77], accumulating localization data can be a major privacy concern [69]. Instead, operational applications need to conceive approaches that only retain insights on locations to shutdown but not individual data. Similarly, any operational use needs to employ differential privacy to limit what stakeholders can learn from the data [4] (e.g., decision-makers can only get a list of candidate locations to close). In the SI Discussion, we further detail approaches to reconcile privacy, ethics and legal considerations.

### Limitations and Future Work

For future investigations of better closure policies, researchers and policymakers need to be cognizant of the limitations of our work. Our analyses capture heterogeneity in individual behavior but does not account for differences in intrinsic vulnerabilities, which are related to severity of risk [35, 55, 23] and disparity in burden of shutdowns on demographic groups [11]. WiMob can be extended with other streams of data to characterize sub-contexts in the population and devise new forms of LC to explicitly study the impact of policies on specific vulnerable subgroups in the community. Additionally, our work explores the avoidance based behavioral responses to closure interventions with assumptions in line with prior work [72, 5]. Researchers and policymakers can be interested in substitution behaviors where the population visits new locations when others are closed. WiMob has the flexibility to model more nuanced spillover effects. Exploring different ways to remove and reallocate edges in the contact network is interesting future work. Further discussion in SI Limitations and Future Work.

## Methods

This section summarizes (i) the data used to derive contact networks and policies, and (ii) the dynamics of our simulation and calibration approach. Additional information for every subsection is present in SI Methods.

### WiFi Mobility

Here we describe the data for our methodology, WiFi mobility models (WiMob) and the process to yield Localized (LC) policies.

#### Data Use and Access

The IT management facility at Georgia Tech (GT) accumulates WiFi access point logs over time. This is common in most universities with managed WiFi infrastructure. We actively collaborated with IT management to define safety and security safeguards that allow us to obtain a de-identified version of these raw logs. Before accessing the data we established a data-use agreement and an ethics protocol that was approved by the Institutional Review Board (IRB) at Georgia Institute of Technology (Protocol H20208). For the WiFi data, we were provided access to logs from Fall 2019 and Fall 2020. We processed these logs to characterize mobility (WiMob) and it encompasses all 40, 000 unique visitors that connected to the network via 6, 959 different access points [15]. The logs did not contain any personally

identifiable information and locations are also coded. The logs indicated the WiFi access point (AP) a device associates with and can therefore be used to infer dwelling locations of users across the entire campus. This is limited to indoor spaces where APs are located and the scope of this localization is at the granularity of a room or suite [20, 76]). For En we only used aggregate insights for enrollment, which were derived from course registration transcripts. Note, we did not cross-identify any students. We used publicly accessible course schedules to approximate schedules of de-identified nodes and infer if they were students or staff, and non-residential or residential. We elaborate on our data in SI Data.

##### Note

Like most universities, GT’s managed WiFi network is not equipped with any Real-Time Location System (RTLS) [14, 46]. RTLS systems use Received Signal Strength Indicator (RSSI) values from multiple neighboring APs to provide high precise localization of individuals in terms of time and space. However, deploying such systems requires surveying the entire network. Additionally, precision localization raises more privacy concerns. These factors together make it challenging for universities to justify the deployment of RTLS, unlike small retail settings that can monetize RTLS insights directly (e.g., insights on footfall can be tied to improving revenue).

#### Contact and Movement Networks

WiMob leverages the logs to create bipartite graphs *K_t_*, for each day *t*, which connect *P* users to *L* access point locations (Figure 1a). Any edge, *p, l _i_* indicates the *i^th^* instance when a *p* was dwelling at *l*. These edges describe the time period of dwelling. Subsequently, by comparing all edges in *K_t_* we can infer if different individuals are collocated near an AP to create a contact network, *G_t_*, for each day *t* — between any collocated *p_i_, p_j_*∈ *P* . These networks define the contact structure for an epidemiological agent-based model at every time-step. Similarly, by inspecting the sequence of dwelling locations for any *p* in graph *K*, we compute a mobility network, *H_t_* — between locations *l* ∈ *L*. In our approach, we considered collocation as a form of *proximate contact* — people in the same room — and therefore established collocation only when this occurred for “an extended period” [30]. By varying this threshold between 30 and 40 minutes we found the contact networks to be structurally similar as their clustering coefficients (over the semester) were highly correlated (*r* = 0.97). In our experiments, we used the 40 minute threshold as it was more computationally less expensive. We provide more details of our approach in SI Data Processing and in SI Modeling Contact and Movement.

#### Modeling Policies

We compared the disease outcomes and burdens of 2 policies, Remote Instruction (RI) and Localized Closure (LC), both of which are modeled with WiMob. For RI we inferred enrollment size of each course in Fall 2019 by determining the number of unique individuals that visited lecture locations during scheduled times. After the first week, we applied the RI by removing all visiting edges in *K_t_* for any *l_c_ L*_RI_ if visits were during lecture times of course *c* with an enrollment ≥ 30. This helped create counterfactual contact networks 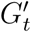. The removal of edges from *K* described the mobility budget of RI and the structure of 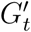 indicated the risk of exposure budget. We designed LC with these budgets by identifying locations for closure (*L*_LC_) with different algorithms, such as *PageRank* [54]*, Eigenvector Centrality* [8]*, Load Centrality* [48]*, and Betweenness Centrality* [24]. When a location was closed, we removed all edges in *K_t_* connected to any *l_x_* ∈ *L*_LC_. We aggregated the movement graph *H_t_* over a week and apply the algorithms to identify locations. Subsequently, we identified the number of top-ranked locations to remove such that the resultant counterfactual contact network 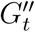 has is within 1% of the budget. The budgets varied for different behavioral scenarios and we only compared policies within the same scenario. This is further elaborated in SI Modeling Policies and Scenarios.

### Disease Simulation

Here we summarize our epidemiological model and calibration process.

#### Agent-Based Model

We constructed an agent-based model (ABM) that captures the spread of COVID-19 between individuals active on campus. This ABM leveraged the contact networks produced by WiMob. The simulation iterated a time-step each day with the underlying contact networks i.e., *G_t_*for no interventions, 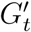 for RI, and 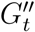 for LC. Each agent in our ABM follows a modified version of *susceptible–exposed–infectious–removed* (SEIR) template that disambiguates the *infectious* compartment into *asymptomatic* and *symptomatic*. New infections were introduced to the model either externally or internally. External transmission arose because individuals could contract the virus outside campus and bring the infection back for local spread [43, 49]. We adopted data of positive cases from Fulton county [51] with a scaling factor *α* to estimate the probability that a *susceptible* individual, who is active on campus, was infected from interactions that take place outside campus. Internal transmissions are determined by *p*, as the probability of *susceptible* individuals in contact with an *infectious* one. We calibrated the parameters related to disease transmission by training and validating our models on the positivity rate reported by GT surveillance testing [28]. SI Agent-Based Model details the disease progression and describes the various parameters.

#### Calibration

In our study, we estimated three key parameters: (i) infectious individuals at day 0, (ii) transmission probability between infectious and susceptible individuals, and (iii) the probability of infection transmission from contacts outside the network. We estimated the range of optimal parameters for disease transmission by minimizing the root means square error (r.m.s.e) between the Georgia Tech surveillance testing positive rates [52, 28] and the observed positivity rate of the model every week— percentage of new *asymptomatic* cases out of the total testable population. The surveillance testing conducted by Georgia Tech was designed for detecting individuals who contracted Covid-19 without showing Flu-like symptoms within the community [52]. We calibrated the model on the positivity rates on the first 5 weeks of Fall 2020. To attain a point estimation of the optimal parameters, we fitted the model to predict trends in the remaining weeks by running a numerical optimization algorithm, Nelder-Mead [44]. To account for quantitative uncertainty, we estimated a range of parameters, within 40% of optimum r.m.s.e [11]. For other model parameters, we adopted values proposed by previous studies on similar populations [36, 27, 42]. Table 2 shows a full list of our parameters.

**Table 2:**
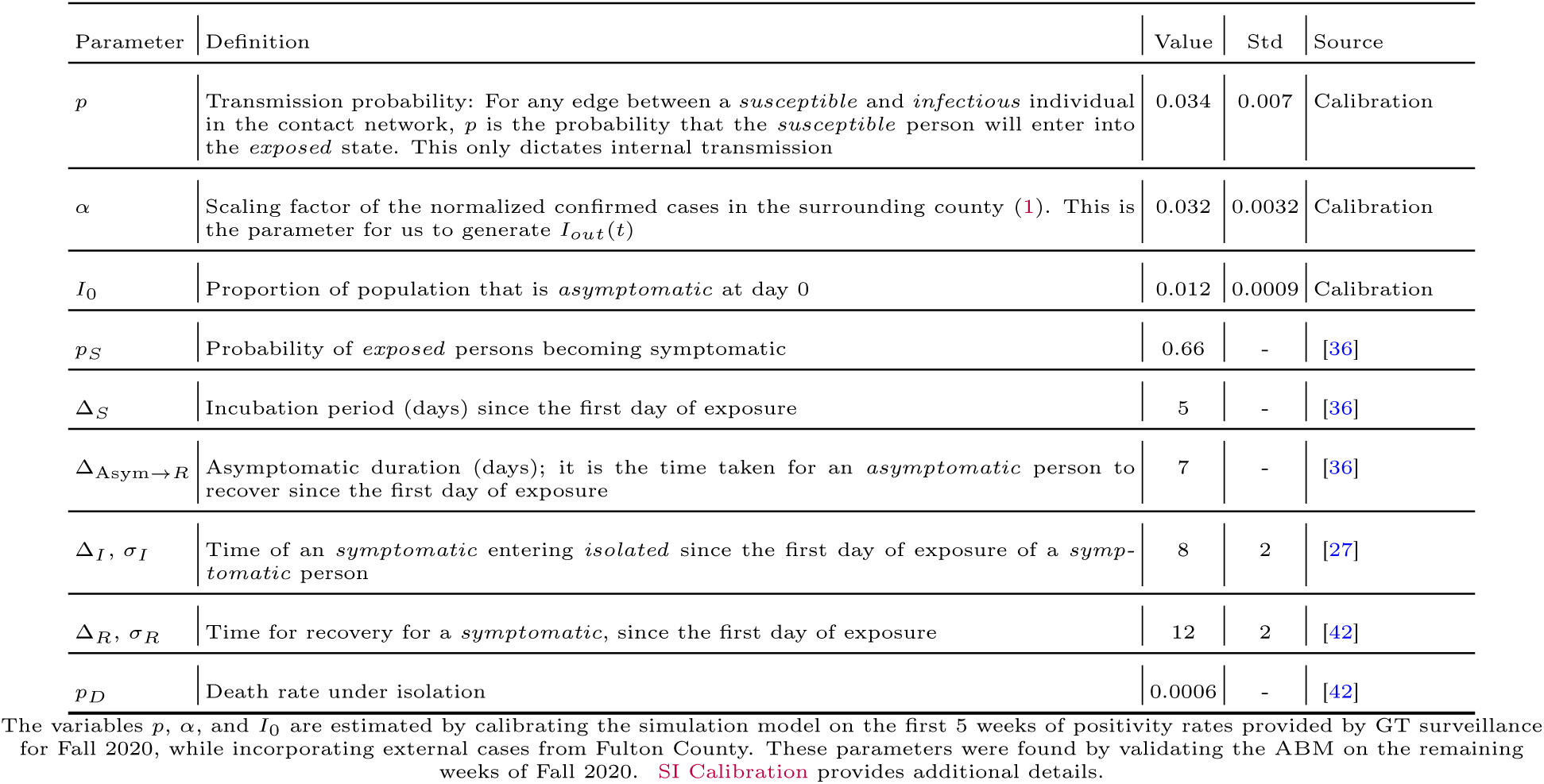
Model Parameters of the ABM

Note that our calibration characterized latent factors associated with pandemic-related cautious behaviors, including the relationship with external transmission. And these factors could be related to “county characteristics, partisanship, media consumption, and racial and ethnic composition” [1]. To account for the effect of these varying latent factors on disease outcomes, we performed additional calibrations for hypothetical variations in disease spread. For these analyses we kept the GT mobility behavior constant while calibrating the model on different time periods of surveillance testing and on positivity rates of different U.S. universities — University of Illinois at Urbana-Champaign [50] and University of California [68], Berkeley. We evaluated RI and LC on these variations and describe the design of these complementary experiments in SI Sensitivity Analyses. See SI Calibration for details on the calibration process and results of all variations are in Table S3.

### Data Availability

Interested parties can request deidentified version of this data through appropriate data use agreements. The data are not publicly available as disclosing them would breach our IRB protocol.

### Code Availability

The code used for processing the WiFi logs into mobility networks can be requested under requisite terms of use agreements. The code for simulation is publicly available at:

https://github.com/AdityaLab/cv-wifi-GT

## Data Availability

Deidentified version of this data can be requested by interested parties through appropriate data use agreements

## Author Contributions Statement

V.D.S., M.D.C., G.D.A., L.N.S., and B.A.P. designed the research; V.D.S., J.X., L.N.S., and B.A.P. performed the research; V.D.S. and G.D.A. acquired the data; V.D.S., J.X., J.C., S.S., and M.M. analyzed, and interpreted data for the work; V.D.S., J.X., M.D.C., G.D.A., L.N.S., and B.A.P. wrote the paper.

## Acknowledgements

This paper is based on work partially supported by the NSF (Expeditions CCF-1918770, CAREER IIS-2028586, RAPID IIS-2027862, RAPID IIS-2027689, Medium IIS-1955883, NRT DGE-1545362, CCF-2115126), CDC MInD program, ORNL, and Semiconductor Research Corporation (in collaboration with Intel Labs). Some research personnel were supported by internal seed funding from the Georgia Institute of Technology and Georgia Tech Research Institute. Other computing resources were provided by the Office of Information Technology at Georgia Tech. The authors thank Di Wu, Hanna Hamilton, and Dima Nazzal (Georgia Institute of Technology) for their analysis of En.

## Supplementary Information Appendix

### Supplementary Methods

In this section, first, we describe the primary data source for mobility models (WiMob), the data used for calibrating our simulations, and for comparison of contact networks with methods using enrollment data (En). Next, we describe how we construct counterfactual mobility networks under the two main policies of interest in our study: remote instruction (RI) and localized closures (LC). Finally, we describe an agent-based-model (ABM) of disease transmission, which has a contact structure based on WiMob, and how this model was calibrated.

#### Data

##### WiFi Mobility

We use data provided by the IT management facility at Georgia Institute of Technology (GT) which accumulates WiFi access point (AP) logs over time. The primary use of WiFi network logs is for maintenance and security purposes. We mine these logs post-hoc to describe the mobility of individuals on campus, which we refer to as WiMob. Here mobility is expressed by visits to certain locations that are demarcated by a corresponding AP. WiMob can also describe dwelling (duration of visits) and collocation (dwelling in the presence of others around the same AP).

The campus WiFi network spans 6959 APs distributed between 240 buildings (and some outdoor locations). We label APs according to which building they are inside, along with the closest room or space (e.g, hallway, lobby, suite, cafe, etc.). The AP may or may not reside inside the room, however, in most cases, only a single AP is associated with space. For less than 5% of the APs, the AP shared association to space with another AP. This many-to-one mapping is typically in the case of large halls and auditoriums. We resolve such many-to-one associations by using APs as a proxy of the space they are associated with. Therefore, individuals connected to different APs in the same space will still be identified as collocated. Similarly, an individual could connect to the network with multiple devices. However, less than 1% logs show that a user is connected to multiple APs around the same time. Therefore, WiMob is agnostic to which device connects to the APs to proxy the presence of the individual. For this study, we obtain the WiFi network logs retrospectively for all of Fall 2019, and the data for Fall 2020 was provided on a per-day basis. Each day, approximately 33, 000 different people connect their devices to the WiFi network on campus. Overall in Fall 2019, approximately 40, 000 different people connected to the campus network.

##### Asymptomatic surveillance testing data

We calibrated the ABM using the publicly reported positivity rate on the GT campus as reported through the asymptomatic surveillance and diagnostic testing program [52]. The testing program used pooled saliva sample surveillance with follow-up diagnostic testing. The positivity rate was reported each day, but individuals must wait at least 1 week between tests. We aggregated the positivity rate by week during the Fall 2020 semester.

##### Confirmed case data

When calibrating our ABM, we considered the reported confirmed cases in Fulton County [51], the county in which GT is located. The ‘Confirmed COVID-19 Cases’ reported in this dataset are cases that have been confirmed with a positive molecular (PCR) test. We considered cases during the Fall 2020 semester to inform external transmissions in the ABM.

##### Enrollment network summary statistics

We compare structural properties of contact networks constructed with WiMob to contact networks constructed from GT’s course enrollment transcripts (En) To ensure that individuals cannot be identified by combining anonymous WiFi network logs and course enrollment transcripts, we only use aggregate statistics from En— structural characteristics of the contact networks described in Table S2. The En network was based on Fall 2019 transcripts for GT’s Atlanta campus. These were cleaned to account for cross-listed courses and was used to determine which students were classmates with each other to form a contact network.

#### WiFi Mobility Models

##### Inferring location from Logs

WiMob is our approach to describe contact between people and movement of people between locations. The first step requires using WiFi network logs to infer when individuals were at specific locations on campus by determining when devices were connected to the corresponding APs. Our system mines the WiFi network logs that are populated via the Simple Network Management Protocol (SNMP) — a standard and widely used monitoring protocol to organize device association behavior to a WiFi network. Periodic SNMP updates can be caused either by poll requests to the APs that log which devices are associated with it at that time. However, devices can appear invisible to detached from an AP for multiple reasons, for example, when devices are idle. Otherwise. SNMP updates can occur whenever a new device connects, which is typical when individuals move between APs. Our approach exploits this factor to first mine periods when individuals are moving, then identify periods of dwelling between movements, and finally determine collocation when two or more individuals are dwelling near the same AP. This system follows from other studies that mine WiFi logs [20, 70] and the detailed processing pipeline and evaluation is presented in [15]. This system to infer collocations has been tested against lecture attendance and reports a high precision of 0.89, but a relatively lower specificity of 0.79 [15]. While it is not likely to show false-positives, it has a possibility to erroneously mark people absent from a location even though they were there. However, for the purposes of our study, a contact network is made over an entire day and it only needs a single collocation instance for us to consider contact. And therefore we believe this limitation would not significantly affect our models.

**Figure S1:**
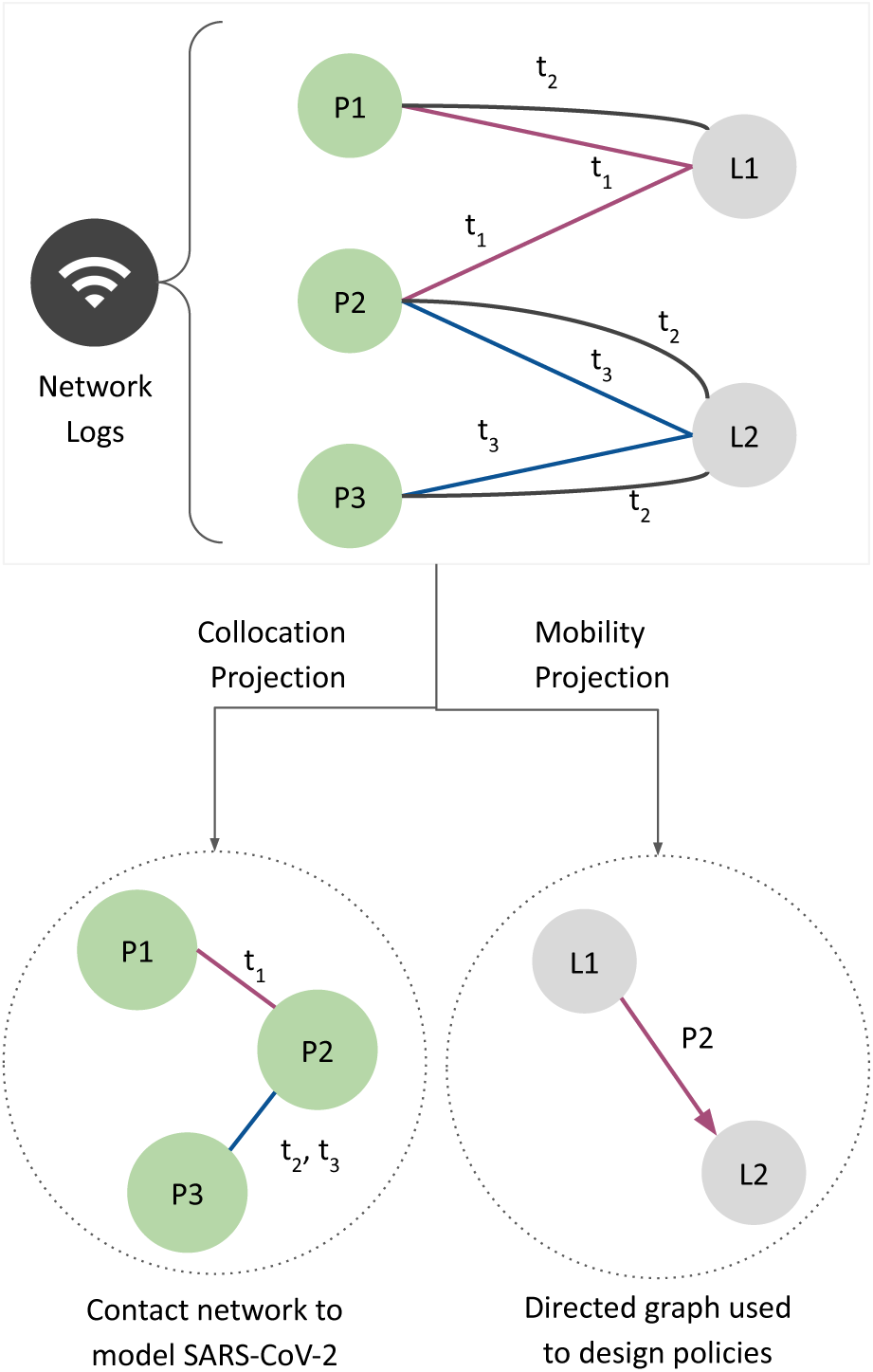
In a managed network, SNMP updates the logs by describing device association to an AP at a certain timestamp. WiMob mines these logs to characterize mobility as a bipartite graph. The nodes are partitioned to describe people nodes (e.g., *P* 1, *P* 2) connected to locations nodes (e.g., *L*1, *L*2). Every edge across the partition describes people visiting locations on campus during different times (e.g., *t*_1_, *t*_2_). Projecting the bipartite on people nodes helps construct a contact network (e.g., *P* 1 and *P* 2 were collocated at *L*1 at *t*_1_), while projecting it on locations helps construct a directed movement graph (*P* 2 dwelled at *L*1 and then at *L*2).

##### Characterizing Logs as Contact and Movement Networks

After inferring where an individual is located on campus, we represent the entire community behavior as graphs. We describe a bipartite graph, *K*, that shows when a user is at a given location on campus (Figure S1). This bipartite graph has edges connecting a set of *m* people, *P* , to a set of *n* locations, *L*. An individual can have multiple edges connecting to the location if they visited that location multiple times (e.g., *t*_1_, *t*_2_). The edge data contains the start and end times of these dwelling periods. For these bipartite graphs, we make a projection on set *P* to describe collocation. This projection graph, *G*, contains an edge between users if they were visiting the same location during overlapping times. Since we do not use RTLS, our approach can only identify if people were in the vicinity of the same AP, but does not describe the distance between them. However, it can reasonably determine collocation in the same room [15]. Since our study is limited to localizing people indoors, we adapt the definition of *proximate contact* [30] where people might be “more than 6 feet but in the same room for an extended period”. In our work, we use a lower bound threshold of 40 minutes to determine proximate contact. Therefore, individuals are only considered in contact when they are collocated in a room for 40 minutes or more. This threshold was set up to account for typical lecture duration on campus (for standard 3-credit hour courses taught 3 times a week). Additionally, we compared the clustering coefficient of the contact networks for different days by varying contact thresholds as 30 and 40 minutes. The Pearson’s r correlation of these was very high 0.97. Thus, we chose to use the 40 minute threshold as it produced structurally similar graphs while requiring lower space constraints. Every edge between two individuals contains a list of locations where they were possibly in contact. *G* forms the basis of the contact-network that we use an agent-based model to simulate. Alternatively, we also make a projection on the set *L*. This projection is a directed graph, *H*, where an edge from *L_i_* to *L_j_* represents movement from the first location to the next within a span of 60 minutes. GT’s large urban campus with pedestrian pathways and motorized transit services enables direct movement between any two places on campus within the threshold. The 60 minutes threshold helps discount erroneously labeling returning from outside campus (e.g., non-residential students visiting two different locations between 2 days). *H* effectively describes how locations are connected and which locations could be more conducive to attracting and disseminating the virus. As a consequence, the *H* helps inform policy design. We compute the bipartite graph and its projections for each day of the semester.

#### Modeling Policies and Scenarios

##### RI: Offering Large Classes Online

As a response to COVID-19, prior work has recommended using En to enforce a form of RI— moving classes large to an online remote instruction setup while other classes are offered in–person [29, 9, 72]. While we have access to aggregate insights on En contact networks, our study protocol prohibits us from accessing course-specific information at an individual level. Therefore to infer individual enrollment, we analyze the edges of the bipartite graph *K*. For this, we first scrape the GT’s course roster for Fall 2019 (filtered to only represent the Atlanta campus). This process provides us with a location and weekly schedule for every lecture conducted on campus, including its various sections. With this information, we are able to identify which edges represent visits to lectures, and subsequently, we can account for unique visitors to a lecture. Thus, we can first identify the number of unique individuals on campus who are enrolled in classes. The aggregate data from course enrollment reports that 21, 299 students were enrolled in Fall 2019. In comparison, our inference identifies 22, 248 students. The excess number can be explained by the fact that our method does not distinguish between instructors, TAs, and students. Next, we study the unique visitors to every lecture in the scraped course schedule which gives us an estimate for the size of every class. Given the limitations of our data processing, actual enrollment sizes could be larger, but our process is less likely to count false positives [15]. Finally, to model RI, for the contact network *G_t_*, we create a counterfactual network 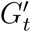 for each day *t*. These exclude collocations that took place at lecture locations during lecture times. If two people were connected solely by proximity during lectures — in a class with large enrollment — they will appear disconnected in the counterfactual network.

##### LC: Closing Important Locations

This article demonstrates the effectiveness of localized closures,LC, which are targeted interventions to seize mobility at different spaces on campus. For this, we identify important locations on campus by analyzing *H*. In the main paper, LC uses *PageRank* [54] as an illustrative algorithm to identify important location nodes. For robustness, we apply various additional algorithms to identify highly authoritative nodes in *H* — betweenness centrality [24], eigenvector centrality [8], and load centrality [48]. In the SI Appendix, we distinguish these different policies as LC_PRank_, LC_BCen_, LC_ECen_, LC_LCen_. Since RI captures a weekly schedule to determine enrollment, LC is implemented to find locations based on behavior from the past 7 days of mobility. We apply the weighted version of the algorithms mentioned earlier on the directed graph representing movement, *H*. The edge weight is based on the number of instances of movement between any *L_i_* and *L_j_* . After sorting the locations by importance, we determine the number of locations to shut down based on different budgets induced by RI— mobility and risk of exposure. For this purpose, we take the approach of a greedy algorithm which successively removes highly-ranked locations till the constraint is met (within 1% margin of error). Similar to RI, LC also render counterfactual collocation networks, *G*”*_t_* for each day *t*. In these networks, we remove instances of collocations that occurred at the shutdown locations. Figure S19 and Figure S20 shows the categories of buildings where different spaces are closed by LC policies.

##### Inducing Budgets and Characterizing Behavioral Scenarios

We now describe how we compare the RI and LC policies. First, we consider the effects of these policies under three behavioral scenarios. These scenarios express the spillover effects of closure that lead to students avoiding campus entirely because their entire schedule is forced online. This analysis assumes that the motivation to be present on campus is determined primarily by enrollment. We consider that, if a student has a full course load (enrolled in a minimum of 3 classes) and all their classes are offered online, that student might have less incentive to visit campus at all (for any engagement) and thus practice *Avoidance*. Since LC could end up closing classrooms, it can also lead to academic schedules being affected and elicit *Avoidance* behavior. As a result, we describe three behavioral scenarios. *Persistence*, is the preliminary, or null scenario, which represents no *Avoidance*. This counterfactual collocation graph only removes edges directly affected by RI or LC. The second scenario we model is *Non-Residential Avoidance* where only non-residential students with full online schedules stop visiting campus entirely. Here the counterfactual graph will remove all edges of non-residential students with fully online schedules. Lastly, the third scenario we model is *Complete Avoidance* where any student with fully online schedules stops activity on campus entirely (including residential students). Here the counterfactual graph will remove all edges from any student with fully online schedules. Since our study protocol prohibits us from mapping our data to other sources, we heuristically infer which individuals are likely to be residential and which are not. We label individuals as residential when they dwell an average of at least 15 minutes at residential locations between 6pm and 10am, on workdays (Monday–Thursday).

Under each behavioral scenario, we limit the number of locations that can be closed under the LC policy to ensure the level of restriction is constrained to be similar to the RI policy. We limit the number of locations under two types of restrictive budgets. The first budget is based on *mobility*, which is the percentage of edges remaining in the bipartite graph if a policy were to be implemented. The second budget is based on *exposure risk*, which is the number of unique individuals who would be in the 1-hop collocation neighborhood of positive individuals. We compute this budget by randomly sampling 2.5% of the population as positive, based on the highest 7-day average positivity rate reported by GT [28] in Fall 2019. Note, however, the effect of RI on campus can vary in different behavioral scenarios, thereby changing the budget available to design a comparable LC policy. For instance, the number of people at exposure risk is much lower in *Complete Avoidance*. As a result, we build multiple alternate networks representing the effect of policies under counterfactual behavioral scenarios.

The infection reduction outcomes and burdens of different policy interventions (under various behavioral scenarios and budgets) is described in Table S4—Table S7 presents box-plots that compares the distribution of disease control outcomes. Figure S11—Figure S14 show cumulative plots of disease control outcomes

#### Agent-based Model

##### Agent-Based Model

We constructed an agent-based model (ABM) that captures the spread of COVID-19 between individuals active within the GT community. The model is used to evaluate the effectiveness of different policy interventions. We consider a modified version of the SEIR framework for simulating the spread of COVID-19 [34, 11] by using an underlying contact network given by WiMob. Figure S2 shows the compartments of the framework. The *susceptible* state (S) represents individuals who have not been infected and can contract the disease by having contact with an infectious individual. The *exposed* state (E) is canonically equivalent to the “incubation period” and is similar to the pre-symptomatic state found in related work [39, 36]. Individuals are considered *infectious* when they are in either the *asymptomatic* state (Asym) or *symptomatic* state (Sym). Individuals in the *asymptomatic* state are assumed to be the major “spreaders” [36] and transmit the infections to *susceptible* individuals before they are *recovered* (R) [23] — after 7 days [36]. Since *asymptomatic* is considered a state of mild severity [32], individuals in this state do not have a risk of fatality. By contrast, for individuals in the *symptomatic* state, will be eventually *isolated* (Iso) (e.g. self-quarantine, or hospitalization on campus). Once in the *isolated* state, they cannot transmit the disease to individuals in the *susceptible* state. Unlike the *asymptomatic* track, the *symptomatic* state is considered critical severity. Therefore, after moving to the *isolated* state, individuals have risk of fatality and entering the death state (D). If the *isolated* individual survives, they enter the *recovered* state. We assume immunity is preserved and therefore after recovery the individual is no longer *susceptible*.

**Figure S2:**
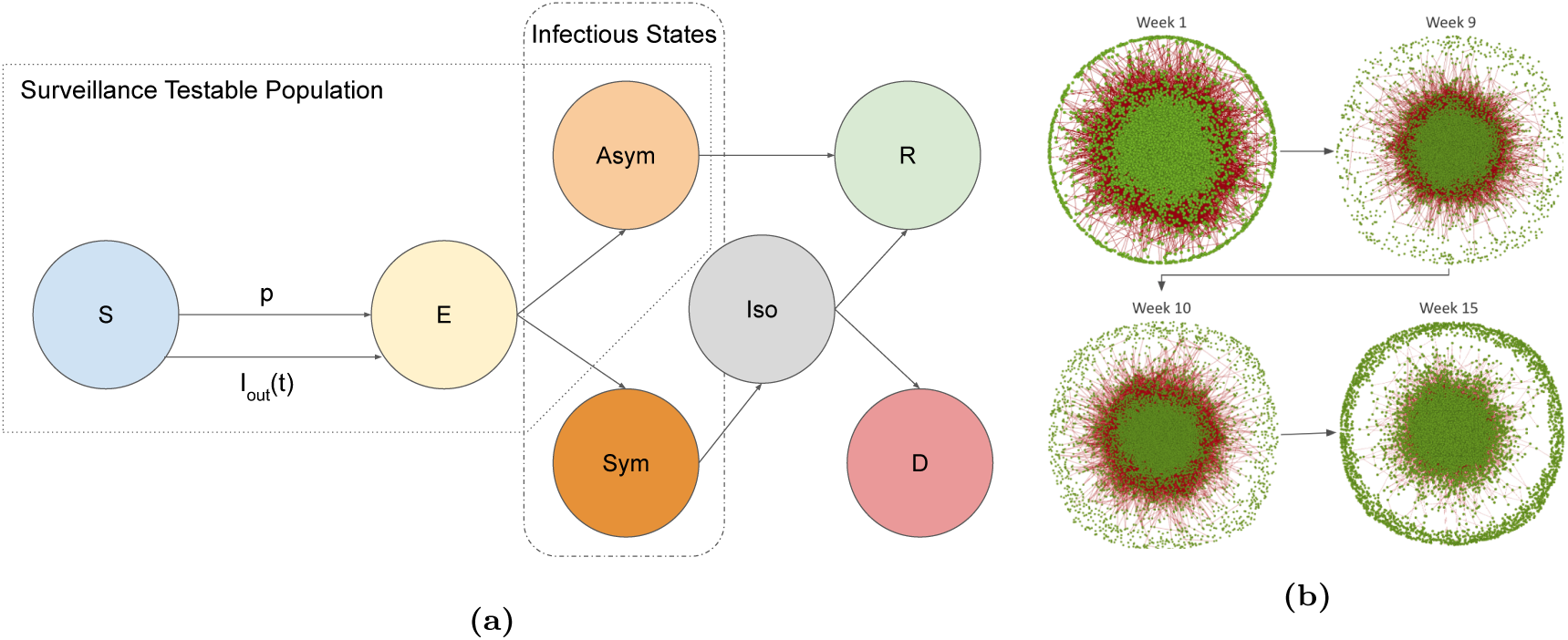
(a) The schematic of the compartments in our modified SEIR model. By the design of the GT surveillance testing [52, 28], the total testable population is defined as the summation of *susceptible*, *exposed*, and *asymptomatic*. Infectious persons are in either *symptomatic* or *symptomatic*. For every effective edge in the mobility network, a susceptible individual that is exposed to an infectious person becomes infected with probability *p*. Individuals may also get infected due to an exposure not captured by the WiMob network which occurs with probability *I_out_*(*t*) on day *t*. account for new infected cases. (b) The mobility behavior represented by WiMob changes every day of the semester (shown weekly here). The contact network constructed from WiMob forms the underlying contact structure of the ABM.

##### Definitions

Let *t* = *{*0, 1, 2, 3*, …, T}* be the index of days in simulations. We denote the sequence of dynamic collocation networks indexed by day *t*, as 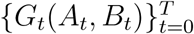 is the set of vertices, individuals on campus, and *B_t_* is the set of edges. The universe set of the population throughout the simulation time period is given by 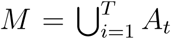. For convenience, we use *a_i_ ∈ M* to index every person in the universe population set.

The SEIR model consists of seven compartments. Each of these corresponds to a function of population subsets with respect to day *t*: *susceptible* S(t), *exposed* E(t), *asymptomatic Asym*(t), *symptomatic Sym*(t), *isolation* I(t), *recovered* R(t), and *dead* D(t). For example, *a_i_ ∈ I*(*t*) means *a_i_* is in the *isolation* state at day *t*. We use 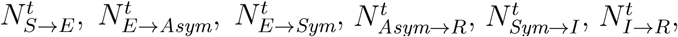 and 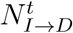 to denote the transitions between states between day *t* and day *t* + 1.

##### Model Initialization

The entire population *M* is fixed where *M* = *S*(*t*)+*E*(*t*)+*Asym*(*t*)+*Sym*(*t*)+*I*(*t*)+*R*(*t*)+*D*(*t*) for all *t*. To capture the positivity out of the students coming back to campus at the start of the semester, we initialize the system by setting a subset of *M* into *Asym*(0) and the reminder into *S*(0). The initial percentage of *asymptomatic* is described by:

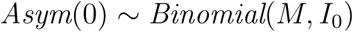

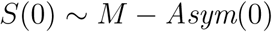

where *I*_0_ is a parameter defined as the initial percentage of *Asymptomatic* at day *t* = 0.

##### New exposures

We consider two ways that an individual in the ABM could be exposed: (i) exposures that occur due to contacts among individuals captured by the mobility network (*internal transmission*) and (ii) exposures that occur due to contacts that occur outside of the mobility network (*external transmission*).

Internal transmissions happen exclusively among individuals in the model. On any given day, an edge becomes effective, when one of the *susceptible* individual comes in contact with the other which is infectious, i.e. *asymptomatic* or *symptomatic*, individual. Therefore, for every effective edge between two such people, the probability of the susceptible individual getting *exposed* is described by the transmission probability *p*, which is another model parameter. The probability for an susceptible individual *a_i_* entering *exposed* at the end of day *t* is given by the following function:

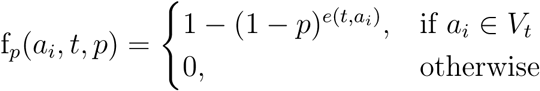

Here, *e*(*t, a_i_*) is the number of effective edges of individual *a_i_*at time *t*. Since (1 − *p*)*^e^*^(^*^t,ai^*^)^ is the probability that *a_i_* does not contracted the disease at time *t* under *e*(*t, a_i_*) Bernoulli trials, 1 (1 *p*)*^e^*^(^*^t,ai^*^)^is the probability that at least one effective edge leading *a_i_*to *exposed*.

In addition to exposure due to internal transmission, we also consider new exposure due to external transmission. We consider external transmission to be exposure resulting from the physical collocations outside the scope of mobility network. For instance, the WiMob does not capture the connections between individuals without access to the campus WiFi or someone contacting infectious persons outside the campus. To reflect this risk in our model, for any day *t*, *I_out_*(*t*) describes the probability of infection on day *t* from a collocation that is external to the mobility network. We assume that the probability an individual is infected due to an external source is proportional to the number of cases in the broader community. Therefore, we model the probability of external infection as a function of confirmed cases in Fulton county, where GT is located [51]. *C_t_* represents the confirmed cases reported by Fulton County where *C_max_* is the maximum number of the cases over the whole period, *I_out_*(*t*) is given by

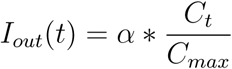

where *α* is a parameter scaling the normalized confirm cases in the surrounding county. The resulting number of external infections on day *t* is then modeled to be are Binomial with *S*(*t*) trials with probability of success *I_out_*(*t*).

In summary, for every day *t >* 0, the overall number of individuals that become newly exposed is represented as 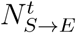 which is the result of both external and internal transmissions.

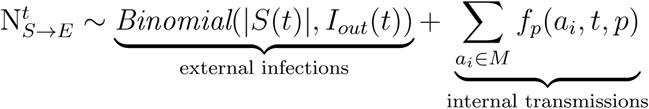

##### Model dynamics after exposure

After exposure, individuals in the model will progress through other disease states in our model. We update the number of individuals in each state daily to reflect transitions between them. The transitions between the states on day *t* are summarized according to the following equations:

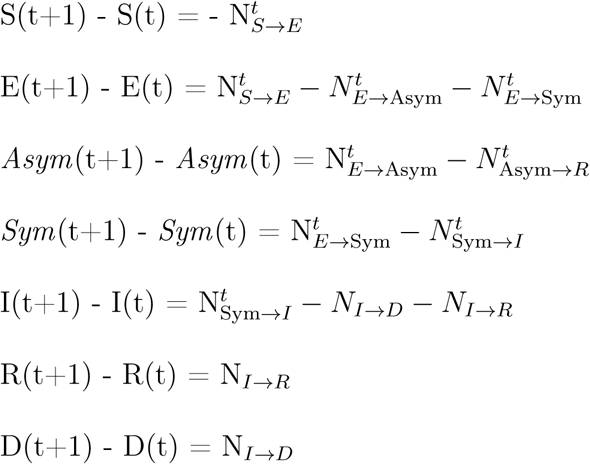

After an individual has been exposed, they will spend Δ_S_ days in an incubation period. At day Δ_S_ after their exposure, individuals will become a *symptomatic* infection with probability *p_S_* . Otherwise the agent will become an *asymptomatic* infection This process is given by the following two equations:

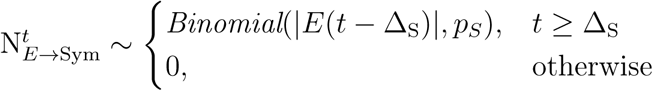

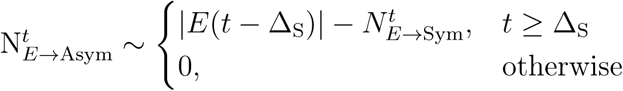

Individuals who enter the *asymptomatic* state will recover after Δ_Asym_*_→R_* days since they were first *exposed*. Thus, we represent the number of transitions from *asymptomatic* to *recovered* on day *t* as:

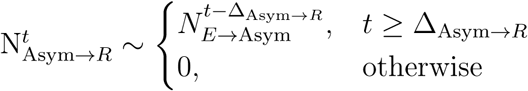

On the other hand, individuals who enter the *symptomatic* will eventually enter the *isolation* state [36]. The time that individuals spend in the *symptomatic* state before entering the *isolated* state is normally distributed 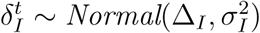. We simulate each individual’s transition between *symptomatic* and *isolated* by using a sampling function Γ(*a_i_, t,* Δ*_t_*) and a function *τ* (*a_i_, t*) that returns the days since *exposed* respectively:

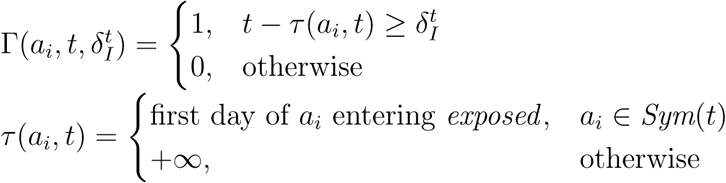

The aggregated transitions 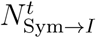 distribution above on each day *t*. between *symptomatic* and *isolated* is the sum of the *t*

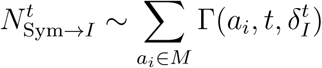

Individuals who enter the *isolated* state may end up with one of two states: *dead* or *recovered*. We defined 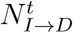 as following another binomial distribution with parameter *p_D_*:

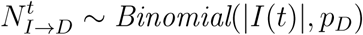

The transitions between *isolation* and *recovered* is quite similar to the transitions between *symptomatic* and *isolation* except 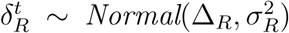 where Δ*_R_* and *σ_R_* are the two parameters standing for the mean and standard deviation of days for an individual in the *isolation* state entering *recovered* since the first day of infection. This leads to:

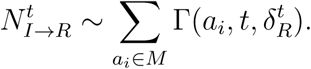

**Table S1:**
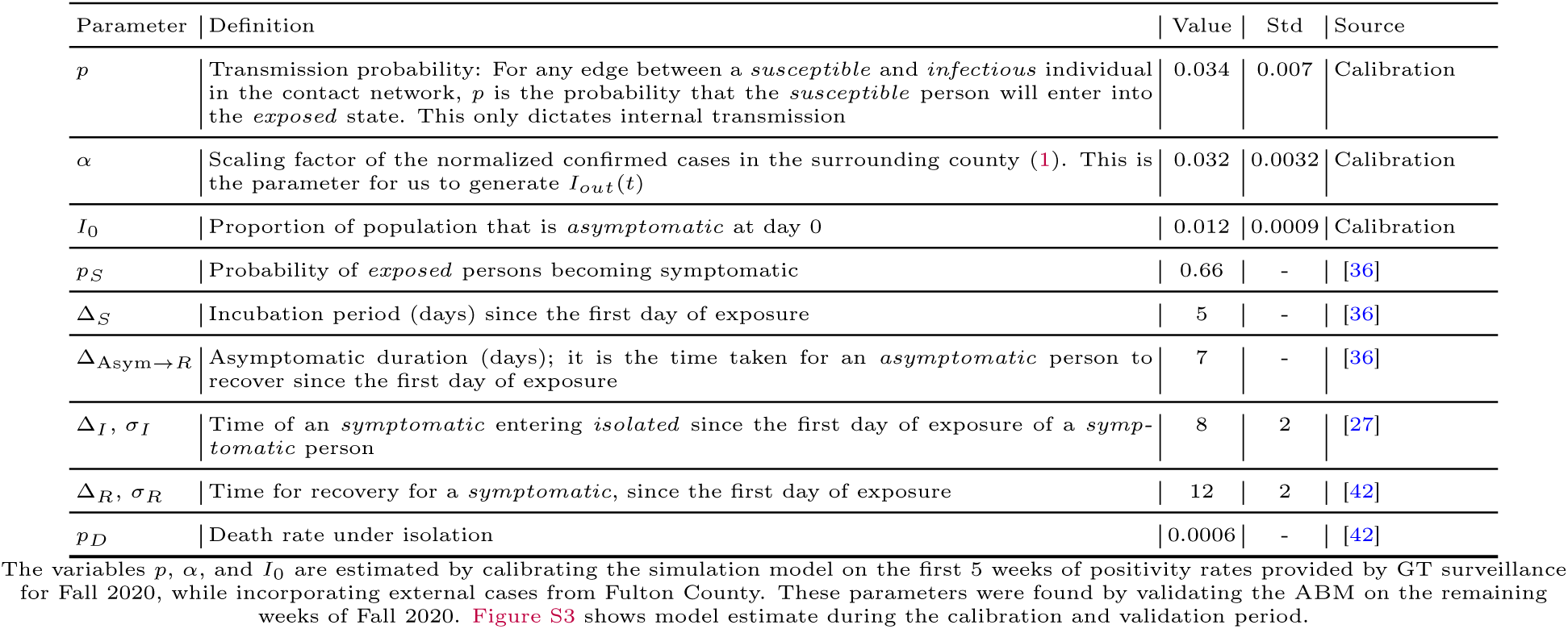
Model Parameters of the ABM

##### Model calibration

Most of our model parameters can be estimated from previous studies (see Table S1). However, three parameters in our study are not easily estimated from previous studies: (i) the proportion of the agents that begin the semester asymptotically infected, *I*_0_, (ii) the probability of transmission between a given infectious individual and susceptible individual given a contact in the mobility network, *p*, and (iii) the scaling factor *α* used to determine probability of transmission due to contact outside of WiMob network on day *t*, *I_out_*(*t*) (see (1)). We fit these three parameters to the published weekly positivity rate (percentage of asymptomatic cases) as reported by GT’s asymptomatic surveillance testing program [52]. To fit the parameters, we performed calibration to minimize the root mean square of error(r.m.s.e) between the simulation estimates of the weekly positivity rate and the observed weekly positivity rate on GT’s campus of the Fall 2020 semester as reported by the surveillance testing program.

To perform the calibration, we used two sets of public data pertaining to 2020 Fall semester at GT: (i) the confirmed cases in Fulton County [51], and (ii) the aggregated surveillance test positivity rate for each week [52]. The former helps estimate the daily external infection percentage. The latter is the ground truth trajectory we fit our model on. We consider the data aggregated by week because each individual on campus can only get tested once per week. The positivity rate provided by the surveillance testing data can be interpreted as the estimated percentage of new *asymptomatic* cases out of the total testable population which includes *susceptible*, *exposed*, and *asymptomatic* — with an assumption that every testable population get tested at the same rate.

To formalize the calibration problem, let *R_w_* be the surveillance-testing aggregated result at week *w*. Let *S*(*I*_0_*, α, p, w*) be the function of the simulation model which returns the percentage of new *asymptomatic* in week *w* out of the total testable population. For every combination of parameters, the predicted result for each week *w* is estimated by taking the average of *N* simulation outputs. The objective function is:

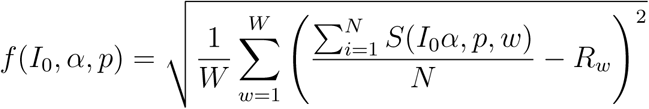

The optimization problem is:

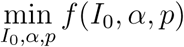

We fit our model to the first 5 weeks of Fall 2020 and validate the results on the remaining weeks. After obtaining the optimal set of parameters, for robust comparison of policies with different viral variants, we generate a range of parameters by compromising the r.m.s.e within 40% of the minima [11]. First, we implement the Nelder Mead method [44] to discover the optimal set of parameters that minimizes the r.m.s.e. Next, we sample 40 different combinations of parameters within 40% of the minimum r.m.s.e to estimate the means and standard deviations of these parameters (Table S1). Throughout this paper, we pool together all simulation results across those parameters over multiple runs (*N* = 15) and report the 2.5th and 97.5th percentiles of the simulation outputs for every policy experiment.

#### Sensitivity Analyses

In this section, we design complementary experiments to inspect the robustness LC policies under different setups and calibration approaches. These variations are defined as follows:

- *Calibration periods (V1)*: For the results in the main paper, we discuss results with our ABM calibrated on the first 5 weeks of surveillance testing data. For additional analyses, the model parameters are re-estimated based on the surveillance data from week 5 9 and 10 14 in Fall 2020 at GT. The calibration is validated on the remaining weeks in the semester. Figure S3 shows the calibration and validation. The results of policy comparison with these variations can be found in Table S8 and Table S9, for weeks 5 9 and 10 14 respectively. Additionally, Figure S9 shows boxplots to compare the distributions of different policies, while Figure S15 and Figure S16 show cumulative plots of the disease control outcomes, for weeks 5*−*9 and 10*−*14 respectively.
- *Campuses and counties (V2)*: For the results in the main paper, the calibration of our ABM reflects certain latent factors inherent to GT that could affect both mobility behavior as well as testing results. To complement this we consider calibrating our data under different settings informed by surveillance testing from other similar large universities. This analysis is intended to represent the GT community in a different geographic setting, which is influenced by a different surrounding community, policies and resources. The new parameters are estimated based on the first 5 weeks of surveillance testing from the University of Illinois at Urbana-Champaign (UIUC) and the University of California, Berkeley (Berkeley) [50, 68], and the corresponding county data [10, 9] The calibration is validated on the remaining weeks in the semester. Figure S4 and Figure S5 show the calibration and validation for UIUC and Berkeley respectively. The results of policy comparison with these variations can be found in Table S10 and Table S11. Additionally, Figure S10 shows boxplots to compare the distributions of different policies, while Figure S17 and Figure S18 show cumulative plots of the disease control outcomes.

The estimated parameters with these calibration variations are described in Table S3. Both RI and LC are evaluated in the same infection reduction metrics and burden metrics again under behavioral scenarios S1, S2, and S3. Since the budgets are structural (mobility, and exposure risk) the LC policies are unchanged among the variants. Moreover, since the burden metrics are structural, those results are invariant.

### Supplementary Discussion

#### Implications for Policy Design

To evaluate the efficacy of policies, we inspect infection reduction by simulating the disease with contact networks from Fall 2019. Since managed WiFi networks accumulate logs for long periods of time, policymakers can use WiMob to model data from previous semesters and experiment with closure policies like LC. We show that WiMob can provide retrospective disease–mitigating insight into multiple counterfactual behavioral scenarios. For instance, policymakers can consider studying seasonal behaviors over multiple semesters for more robustness. Since the underlying data is longitudinal, it provides the flexibility to realistically assess policy interventions at different time points and also study updating policies. Restricting movement on campus at different time-points is known to exert varying degrees of control on disease spread [11]. Our data also shows that mobility on campus varies across the semester and therefore, allows policymakers to consider loosening shutdowns depending on the phase of the semester.

Policy design is determined by practical budgets. We model two kinds of budgets, mobility reduction and risk of exposure. The former represents disruptions in space utilization, availing services, and social life. The latter translates to the testing burden on campus. Our analysis determines the budget in different behavioral scenarios by observing the changes to the graph when large classes are moved online. This is to ensure an equitable comparison with targeted policies. However, in real situations, these budgets can be relaxed or restricted based on that campus’ preparedness to tackle a pandemic. For instance, a hypothetical campus that can test everyone every day might not be constrained by risk of exposure. Alternatively, policymakers can model other tangible budgets such as the capacity in isolation wards or available hospital beds. This can be informed by practical limitations of the campus. Similarly, this paper only assesses limited forms of cost, e.g., students avoiding campus or closing locations. From a financial perspective, university campuses can digitize their core service—education—but still realize losses from other curtailed services [21, 7, 71]. When students avoid campus it can lead to direct losses from meal passes and parking and also quantifiable losses to learning outcomes [2, 18] Policymakers can compute actual costs by complementing this data with information from other sources (e.g., revenue generated by cafes and stores on campus). This can help qualifying WiMob to reflect different costs and in turn help design policies that optimize for financial losses. Different campuses have different priorities and challenges in implementing policies.

#### Privacy, Ethics and Legal Considerations

We purposefully compare our prototype targeted policies against moving classes online because of practical budgets within the university. Both the WiMob and En based contact networks are derived from archival data accumulated by universities. This does not require instrumenting campus or its community with any new form of surveillance infrastructure. However, its use for a different purpose demands approval by an IRB. Moreover, acquiring these kinds of data would require collaborating with data-stewards (e.g., the IT department) to establish a data-use agreement. This document must clarify how the data will be de-identified, transferred, and stored.

For this form of data, the critical privacy challenge might not be localization itself, but rather the aggregation of data over a period of time [69]. Data spanning a longer period are more susceptible to cross-analyzing and identifying. To mitigate over-accumulation of data, we suggest an adherence to principles of data minimization [31]. Instead of storing entire mobility graphs, the campus can compute and preserve only high-level insights, such as the importance of locations. This redacts any underlying individual behavior and corresponding identifiable information. Actually, for future purposes campuses can consider a form of differential privacy that authorizes limited forms of data querying depending on the privileges of the stakeholder [4].

An operational application would require the university to update the terms of use for its managed network. Particularly, the university should disclose how this data can be used in critical circumstances that invoke shared vulnerabilities [6]. On notifying the campus community of this change it offers individuals the choice to refrain from using the university network. Prior work on a sample within the same university campus shows that 90% of students are connected to the network on any given day [15]. Therefore, proposing such an opt-out condition can be viewed as an unfair choice. As a result, the campus needs to develop a contingency plan to accommodate network access to users who do not want their mobility behavior to constitute the aggregated insights.

#### Limitations and Future Work

This work presents evidence that university campuses can repurpose existing data sources to inform the design of LC policies that can control COVID-19. We evaluate these policies as alternatives to other data-driven, but, broad impact policies that universities consider implementing, such as moving large classes online. One of the drawbacks of this analysis, however, is that it assumes all edges to be the same. For example, when constraining by mobility, in real scenarios losing certain visits might be more valuable than others. Decline in mobility around profit-making services, such as shops and cafeterias, versus losing mobility at common rooms have a different tangible effects on campus. Currently, we take an agnostic stance towards the mobility behavior, where all visits at all locations are the same. In reality, implementing policies could have inequitable qualitative impacts despite appearing to have a similar network configuration. This can be improved by embedding more qualitative information into the network and conceiving ingenious ways to associate costs to edges.

Similar to the assumption that all visits and locations, the current work also assumes all people to be equal. However, different people have different underlying conditions that can make their vulnerabilities more concerning [55]. The privacy safeguards of this study restricted the research team from acquiring any additional demographic or historical information. Further work can attempt to characterize the nodes by randomly seeding the network to reflect the approximate demographic break up of the community. Alternatively, researchers could try to estimate some demographic based on behavior as well. However, to leverage accurate individual information, even for operational use during a public health emergency, policymakers and researchers need to develop new privacy protocols [24].

Lastly, this paper only studies three rudimentary behavioral scenarios, *persistence*, *non-residential avoidance*, *complete avoidance*. Yet, other substitution behaviors are possible and the richness of networks leveraged with WiMob enables the exploration of various new scenarios that can be triggered by policy interventions on campus. For instance, individuals might not even visit transitory spaces, such as lobbies or cafes between classes. Certain collocations could be the consequence of social ties which might never be developed because of a shutdown (e.g., project teams meeting outside of class). Further research can illuminate the effects of policies in more specific scenarios by modeling post-intervention behavior more accurately.

**Table S2:**
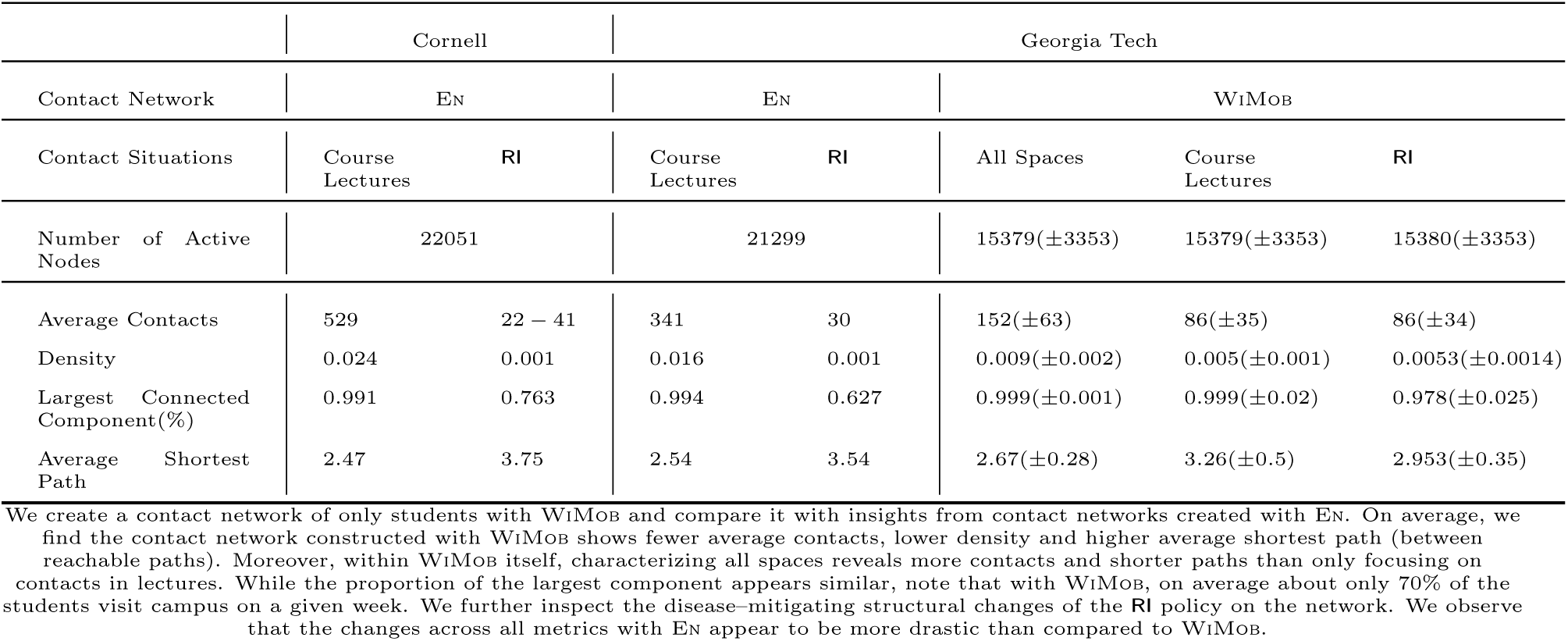
Comparison of Contact Network Structure (Fall 2019)

**Table S3:**
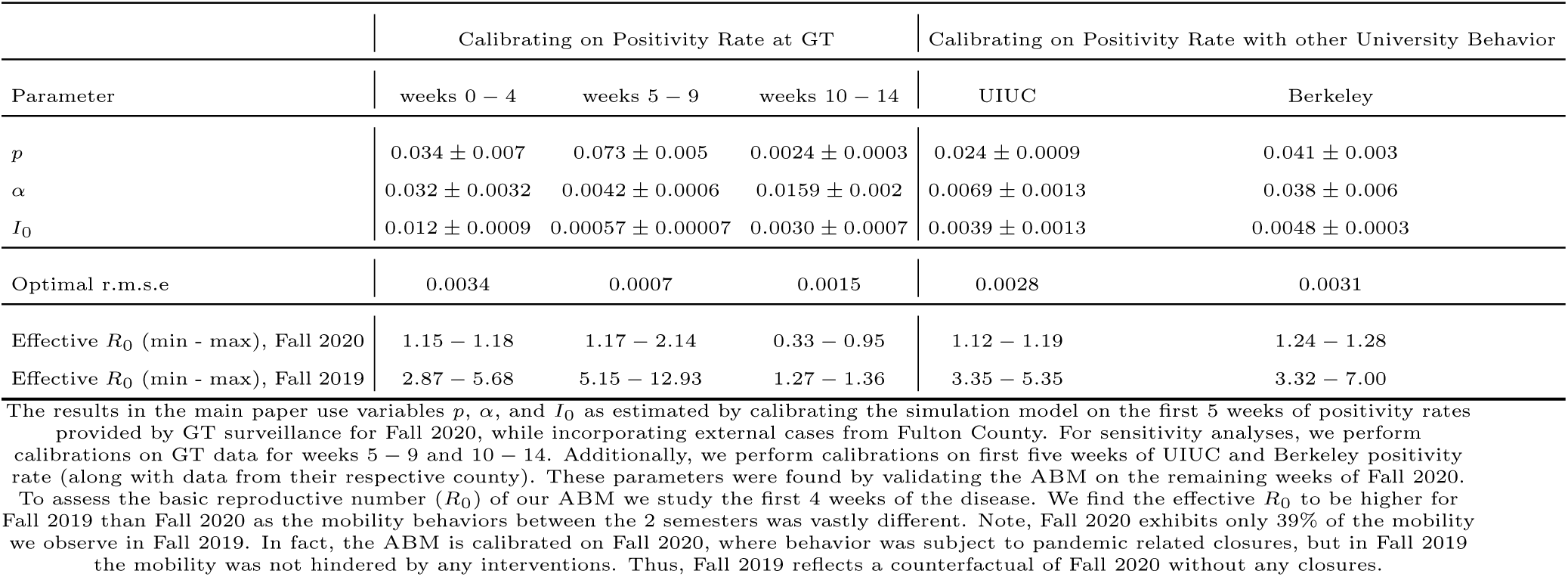
Calibration outcomes with variations

**Table S4:**
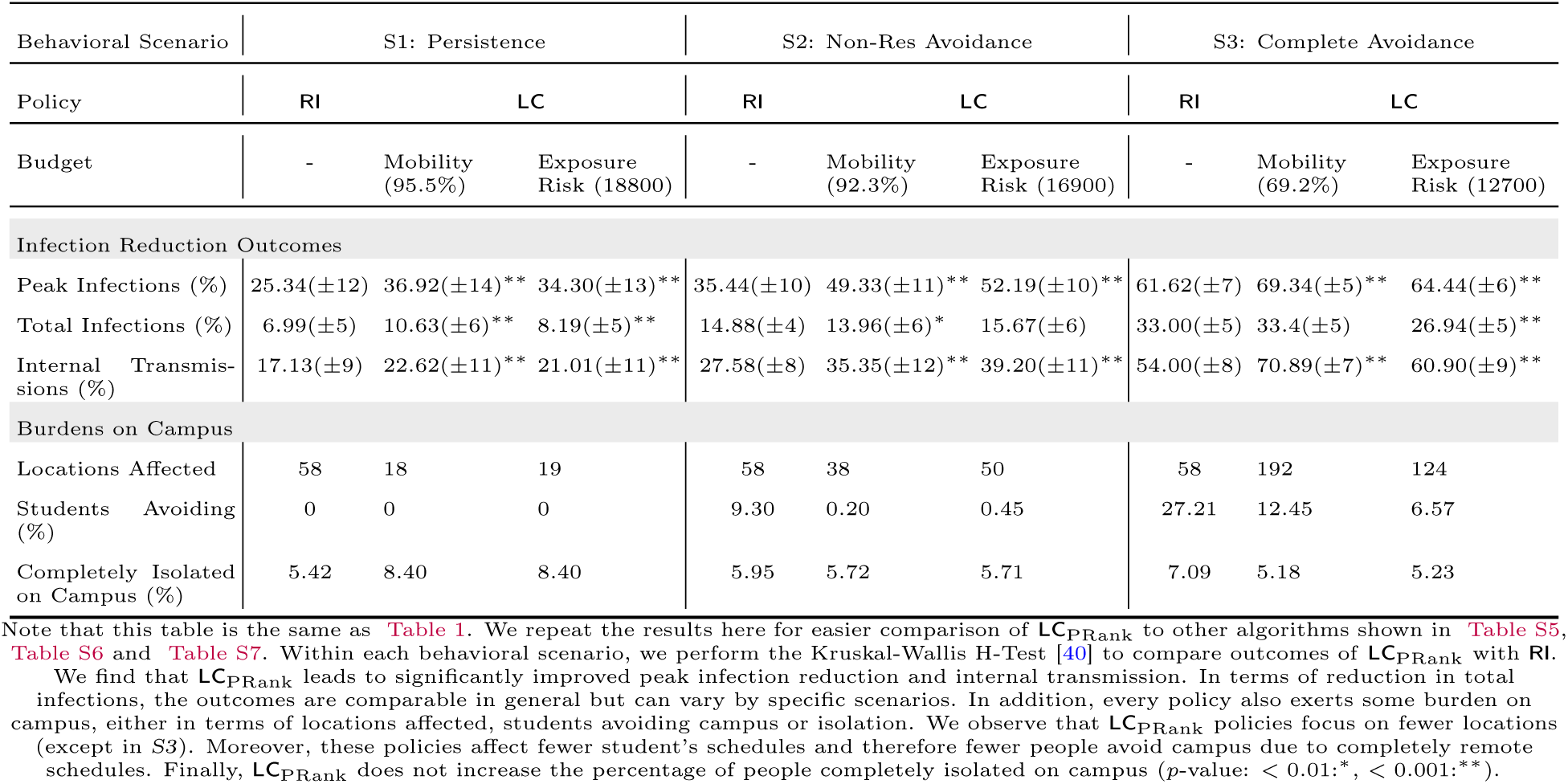
Comparison of different LC_PRank_ policies in terms of controlling the disease and impacts on campus in Fall 2019; calibrated from week 0 *−* 4 in Fall 2020 at GT

**Table S5:**
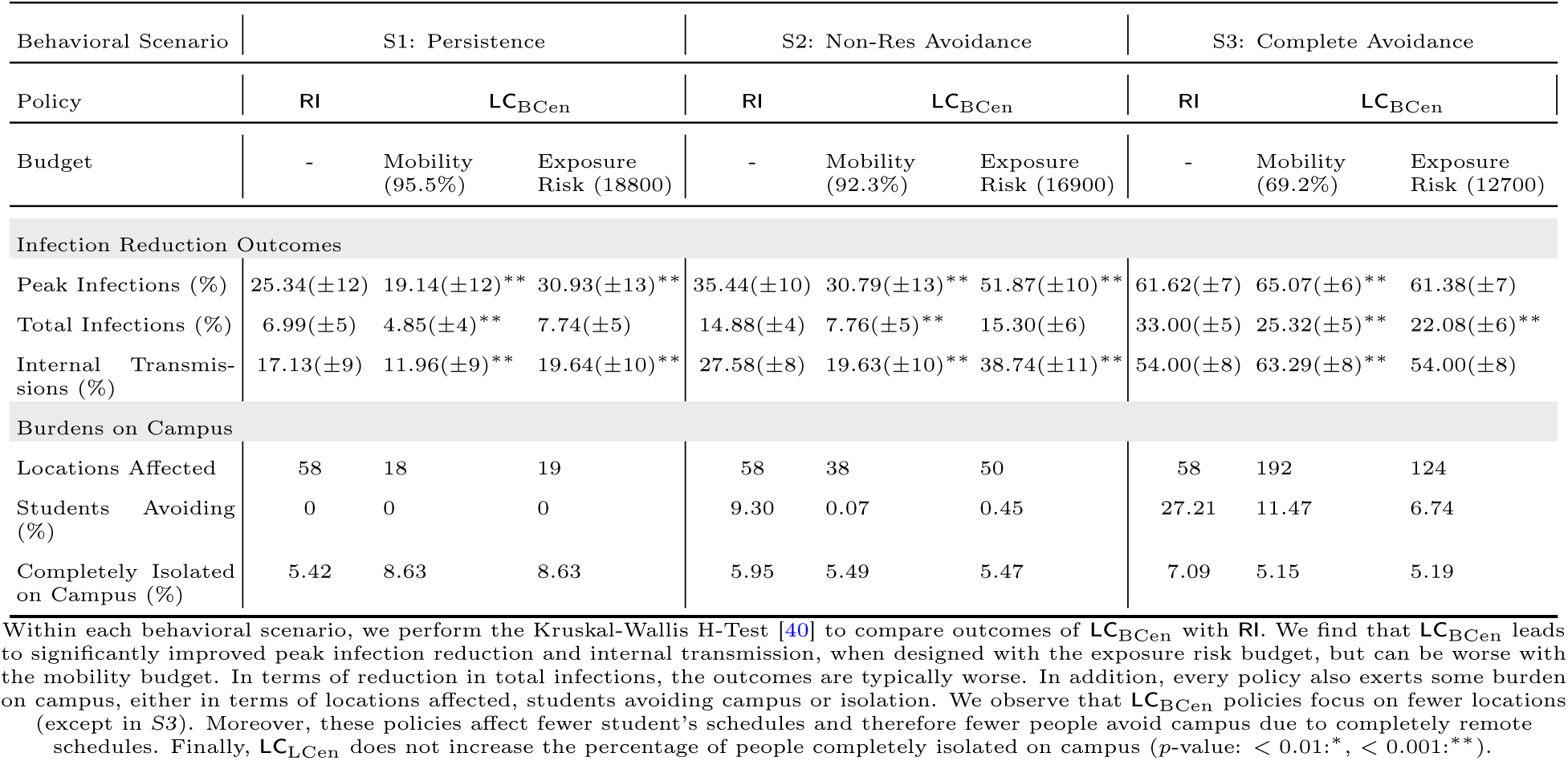
Comparison of different LC_BCen_ policies in terms of controlling the disease and impacts on campus in Fall 2019; calibrated from week 0 − 4 in Fall 2020 at GT

| Behavioral Scenario | S1: Persistence |  |  | S2: Non-Res Avoidance |  |  | S3: Complete Avoidance |  |  |
| --- | --- | --- | --- | --- | --- | --- | --- | --- | --- |
| Policy | RI | $LC_{BCen}$ | | RI | $LC_{BCen}$ | | RI | $LC_{BCen}$ | |
| Budget | - | Mobility<br>(95.5%) | Exposure<br>Risk (18800) | - | Mobility<br>(92.3%) | Exposure<br>Risk (16900) | - | Mobility<br>(69.2%) | Exposure<br>Risk (12700) |
| **Infection Reduction Outcomes** |  |  |  |  |  |  |  |  |  |
| Peak Infections (%) | 25.34(±12) | 19.14(±12)** | 30.93(±13)** | 35.44(±10) | 30.79(±13)** | 51.87(±10)** | 61.62(±7) | 65.07(±6)** | 61.38(±7) |
| Total Infections (%) | 6.99(±5) | 4.85(±4)** | 7.74(±5) | 14.88(±4) | 7.76(±5)** | 15.30(±6) | 33.00(±5) | 25.32(±5)** | 22.08(±6)** |
| Internal Transmissions (%) | 17.13(±9) | 11.96(±9)** | 19.64(±10)** | 27.58(±8) | 19.63(±10)** | 38.74(±11)** | 54.00(±8) | 63.29(±8)** | 54.00(±8) |
| **Burdens on Campus** |  |  |  |  |  |  |  |  |  |
| Locations Affected | 58 | 18 | 19 | 58 | 38 | 50 | 58 | 192 | 124 |
| Students Avoiding (%) | 0 | 0 | 0 | 9.30 | 0.07 | 0.45 | 27.21 | 11.47 | 6.74 |
| Completely Isolated on Campus (%) | 5.42 | 8.63 | 8.63 | 5.95 | 5.49 | 5.47 | 7.09 | 5.15 | 5.19 |

Within each behavioral scenario, we perform the Kruskal-Wallis H-Test [40] to compare outcomes of $LC_{BCen}$ with RI. We find that $LC_{BCen}$ leads to significantly improved peak infection reduction and internal transmission, when designed with the exposure risk budget, but can be worse with the mobility budget. In terms of reduction in total infections, the outcomes are typically worse. In addition, every policy also exerts some burden on campus, either in terms of locations affected, students avoiding campus or isolation. We observe that $LC_{BCen}$ policies focus on fewer locations (except in S3). Moreover, these policies affect fewer student's schedules and therefore fewer people avoid campus due to completely remote schedules. Finally, $LC_{BCen}$ does not increase the percentage of people completely isolated on campus ( $p$ -value: $< 0.01$ \*, $< 0.001$ \*\*).

**Table S6:** Comparison of different LC_ECen_ policies in terms of controlling the disease and impacts on campus in Fall 2019; calibrated from week 0 *−* 4 in Fall 2020 at GT

| Behavioral Scenario | S1: Persistence |  |  | S2: Non-Res Avoidance |  |  | S3: Complete Avoidance |  |  |
| --- | --- | --- | --- | --- | --- | --- | --- | --- | --- |
| Policy | RI | $LC_{ECen}$ | | RI | $LC_{ECen}$ | | RI | $LC_{ECen}$ | |
| Budget | - | Mobility (95.5%) | Exposure Risk (18800) | - | Mobility (92.3%) | Exposure Risk (16900) | - | Mobility (69.2%) | Exposure Risk (12700) |
| **Infection Reduction Outcomes** |  |  |  |  |  |  |  |  |  |
| Peak Infections (%) | 25.34(±12) | 36.15(±13)** | 36.13(±13)** | 35.44(±10) | 44.52(±12)** | 51.33(±10)** | 61.62(±7) | 65.13(±6)** | 62.15(±7) |
| Total Infections (%) | 6.99(±5) | 8.66(±6)** | 8.69(±6)** | 14.88(±4) | 11.75(±6)** | 14.96(±6) | 33.00(±5) | 25.39(±5)** | 22.82(±6)** |
| Internal Transmissions (%) | 17.13(±9) | 22.33(±11)** | 22.37(±11)** | 27.58(±8) | 29.95(±12)* | 37.94(±11)** | 54.00(±8) | 63.56(±8)** | 57.07(±10)** |
| **Burdens on Campus** |  |  |  |  |  |  |  |  |  |
| Locations Affected | 58 | 18 | 19 | 58 | 38 | 50 | 58 | 192 | 124 |
| Students Avoiding (%) | 0 | 0 | 0 | 9.30 | 0.20 | 0.55 | 27.21 | 13.11 | 6.96 |
| Completely Isolated on Campus (%) | 5.42 | 8.59 | 8.59 | 5.95 | 5.53 | 5.51 | 7.09 | 5.17 | 5.23 |

Within each behavioral scenario, we perform the Kruskal-Wallis H-Test [40] to compare outcomes of $LC_{ECen}$ with RI. We find that $LC_{ECen}$ leads to significantly improved peak infection reduction and internal transmission. In terms of reduction in total infections, the outcomes vary by specific scenarios. In addition, every policy also exerts some burden on campus, either in terms of locations affected, students avoiding campus or isolation. We observe that $LC_{ECen}$ policies focus on fewer locations (except in S3). Moreover, these policies affect fewer student's schedules and therefore fewer people avoid campus due to completely remote schedules. Finally, $LC_{ECen}$ does not increase the percentage of people completely isolated on campus ( $p$ -value: $< 0.01$ \*, $< 0.001$ \*\*).

**Table S7:**
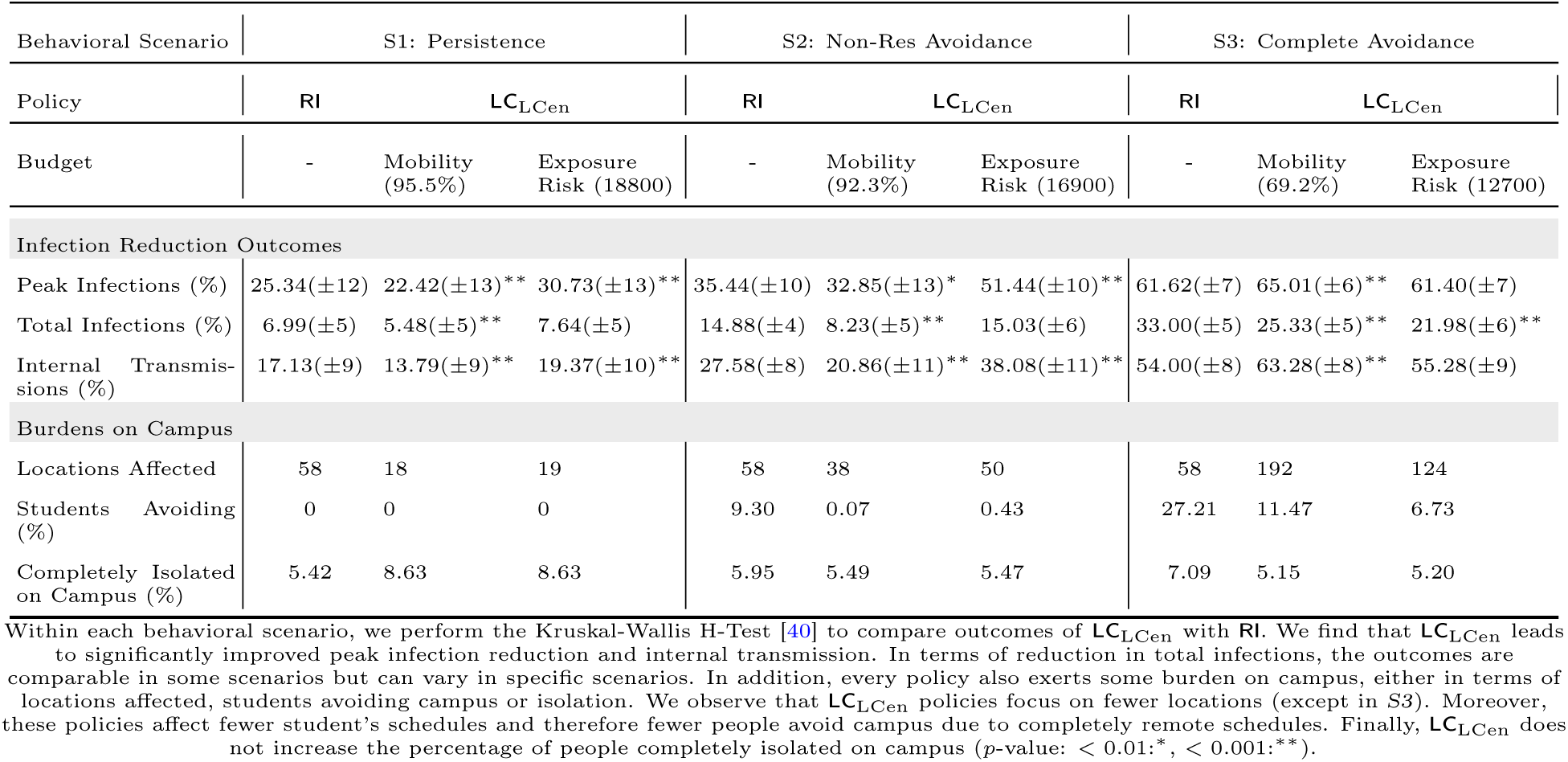
Comparison of different LC_LCen_ policies in terms of controlling the disease and impacts on campus in Fall 2019; calibrated from week 0 − 4 in Fall 2020 at GT

**Table S8:** Comparison of different LC_PRank_ policies in terms of controlling the disease and impacts on campus in Fall 2019; calibrated from week 5 *−* 9 in Fall 2020 at GT

| Behavioral Scenario | S1: Persistence |  |  | S2: Non-Res Avoidance |  |  | S3: Complete Avoidance |  |  |
| --- | --- | --- | --- | --- | --- | --- | --- | --- | --- |
| Policy | Broad | $LC_{PRank}$ | | RI | $LC_{PRank}$ | | RI | $LC_{PRank}$ | |
| Budget | - | Mobility<br>(95.5%) | Exposure<br>Risk (18800) | - | Mobility<br>(92.3%) | Exposure<br>Risk (16900) | - | Mobility<br>(69.2%) | Exposure<br>Risk (12700) |
| Infection Reduction Outcomes |  |  |  |  |  |  |  |  |  |
| Peak Infections (%) | 20.10($\pm 4$) | 25.60($\pm 3$)** | 25.63($\pm 3$)** | 31.25($\pm 3$) | 42.32($\pm 4$)** | 47.29($\pm 4$)** | 62.35($\pm 2$) | 88.87($\pm 2$)** | 76.89($\pm 3$)** |
| Total Infections (%) | 8.89($\pm 2$) | 10.50($\pm 3$)** | 9.70($\pm 3$)** | 20.26($\pm 2$) | 20.02($\pm 3$) | 23.71($\pm 4$)** | 46.72($\pm 2$) | 67.92($\pm 4$)** | 51.30($\pm 4$)** |
| Internal Transmissions (%) | 9.97($\pm 2$) | 11.51($\pm 2$)** | 10.95($\pm 2$)** | 21.84($\pm 2$) | 22.51($\pm 3$) | 26.64($\pm 3$)** | 49.80($\pm 2$) | 74.96($\pm 3$)** | 56.89($\pm 4$)** |

**Table S9:** Comparison of different LC_PRank_ policies in terms of controlling the disease and impacts on campus in Fall 2019; calibrated from week 10 − 14 in Fall 2020 at GT

| Behavioral Scenario | S1: Persistence |  |  | S2: Non-Res Avoidance |  |  | S3: Complete Avoidance |  |  |
| --- | --- | --- | --- | --- | --- | --- | --- | --- | --- |
| Policy | Broad | $LC_{PRank}$ | | RI | $LC_{PRank}$ | | RI | $LC_{PRank}$ | |
| Budget | - | Mobility<br>(95.5%) | Exposure<br>Risk (18800) | - | Mobility<br>(92.3%) | Exposure<br>Risk (16900) | - | Mobility<br>(69.2%) | Exposure<br>Risk (12700) |
| Infection Reduction Outcomes |  |  |  |  |  |  |  |  |  |
| Peak Infections (%) | -1.75($\pm 8$) | 3.65($\pm 8$)** | -1.95($\pm 8$) | 3.88($\pm 8$) | -2.24($\pm 8$)** | -2.06($\pm 8$)** | 20.39($\pm 7$) | 7.57($\pm 8$)** | 2.81($\pm 8$)** |
| Total Infections (%) | 3.93($\pm 9$) | 10.36($\pm 8$)** | 5.13($\pm 9$) | 9.87($\pm 8$) | 6.36($\pm 9$)** | 6.48($\pm 9$)** | 26.02($\pm 7$) | 16.37($\pm 8$)** | 11.80($\pm 8$)** |
| Internal Transmissions (%) | 42.33($\pm 10$) | 61.15($\pm 7$)** | 56.25($\pm 8$)** | 49.83($\pm 9$) | 67.10($\pm 6$)** | 69.10($\pm 6$)** | 74.74($\pm 5$) | 84.80($\pm 3$)** | 79.90($\pm 4$)** |

**Table S10:** Comparison of different LC_PRank_ policies in terms of controlling the disease and impacts on campus in Fall 2019; calibrated from week 0 *−* 4 in Fall 2020 at UIUC

| Behavioral Scenario | S1: Persistence |  |  | S2: Non-Res Avoidance |  |  | S3: Complete Avoidance |  |  |
| --- | --- | --- | --- | --- | --- | --- | --- | --- | --- |
| Policy | Broad | $\text{LC}_{\text{PRank}}$ | | RI | $\text{LC}_{\text{PRank}}$ | | RI | $\text{LC}_{\text{PRank}}$ | |
| Budget | - | Mobility<br>(95.5%) | Exposure<br>Risk (18800) | - | Mobility<br>(92.3%) | Exposure<br>Risk (16900) | - | Mobility<br>(69.2%) | Exposure<br>Risk (12700) |
| **Infection Reduction Outcomes** |  |  |  |  |  |  |  |  |  |
| Peak Infections (%) | 41.40($\pm 3$) | 60.44($\pm 2$)** | 59.52($\pm 2$)** | 49.75($\pm 2$) | 74.22($\pm 2$)** | 76.44($\pm 2$)** | 78.14($\pm 1$) | 85.81($\pm 1$)** | 83.71($\pm 1$)** |
| Total Infections (%) | 18.46($\pm 3$) | 27.12($\pm 3$)** | 25.25($\pm 3$)** | 27.09($\pm 3$) | 38.00($\pm 4$)** | 40.68($\pm 4$)** | 51.97($\pm 3$) | 59.93($\pm 5$)** | 54.07($\pm 5$)** |
| Internal Transmissions (%) | 28.22($\pm 3$) | 40.93($\pm 3$)** | 39.09($\pm 3$)** | 37.89($\pm 3$) | 58.47($\pm 2$)** | 65.45($\pm 2$)** | 68.04($\pm 2$) | 86.45($\pm 1$)** | 80.08($\pm 1$)** |

**Table S11:** Comparison of different LC_PRank_ policies in terms of controlling the disease and impacts on campus in Fall 2019; calibrated from week 0 − 4 in Fall 2020 at UC Berkeley

| Behavioral Scenario | S1: Persistence |  |  | S2: Non-Res Avoidance |  |  | S3: Complete Avoidance |  |  |
| --- | --- | --- | --- | --- | --- | --- | --- | --- | --- |
| Policy | Broad | $\text{LC}_{\text{PRank}}$ | | RI | $\text{LC}_{\text{PRank}}$ | | RI | $\text{LC}_{\text{PRank}}$ | |
| Budget | - | Mobility<br>(95.5%) | Exposure<br>Risk (18800) | - | Mobility<br>(92.3%) | Exposure<br>Risk (16900) | - | Mobility<br>(69.2%) | Exposure<br>Risk (12700) |
| **Infection Reduction Outcomes** |  |  |  |  |  |  |  |  |  |
| Peak Infections (%) | 29.13($\pm 3$) | 36.46($\pm 5$)** | 36.34($\pm 5$)** | 38.83($\pm 3$) | 54.95($\pm 4$)** | 58.88($\pm 4$)** | 66.69($\pm 2$) | 78.18($\pm 1$)** | 77.65($\pm 2$)** |
| Total Infections (%) | 6.34($\pm 3$) | 8.59($\pm 3$)** | 7.28($\pm 3$)** | 14.71($\pm 3$) | 13.18($\pm 4$)** | 14.83($\pm 4$) | 33.86($\pm 4$) | 33.98($\pm 5$) | 27.10($\pm 5$)** |
| Internal Transmissions (%) | 15.99($\pm 3$) | 20.43($\pm 4$)** | 19.17($\pm 4$)** | 27.01($\pm 3$) | 34.60($\pm 4$)** | 38.78($\pm 4$)** | 55.01($\pm 2$) | 74.65($\pm 2$)** | 63.57($\pm 3$)** |

**Figure S3:**
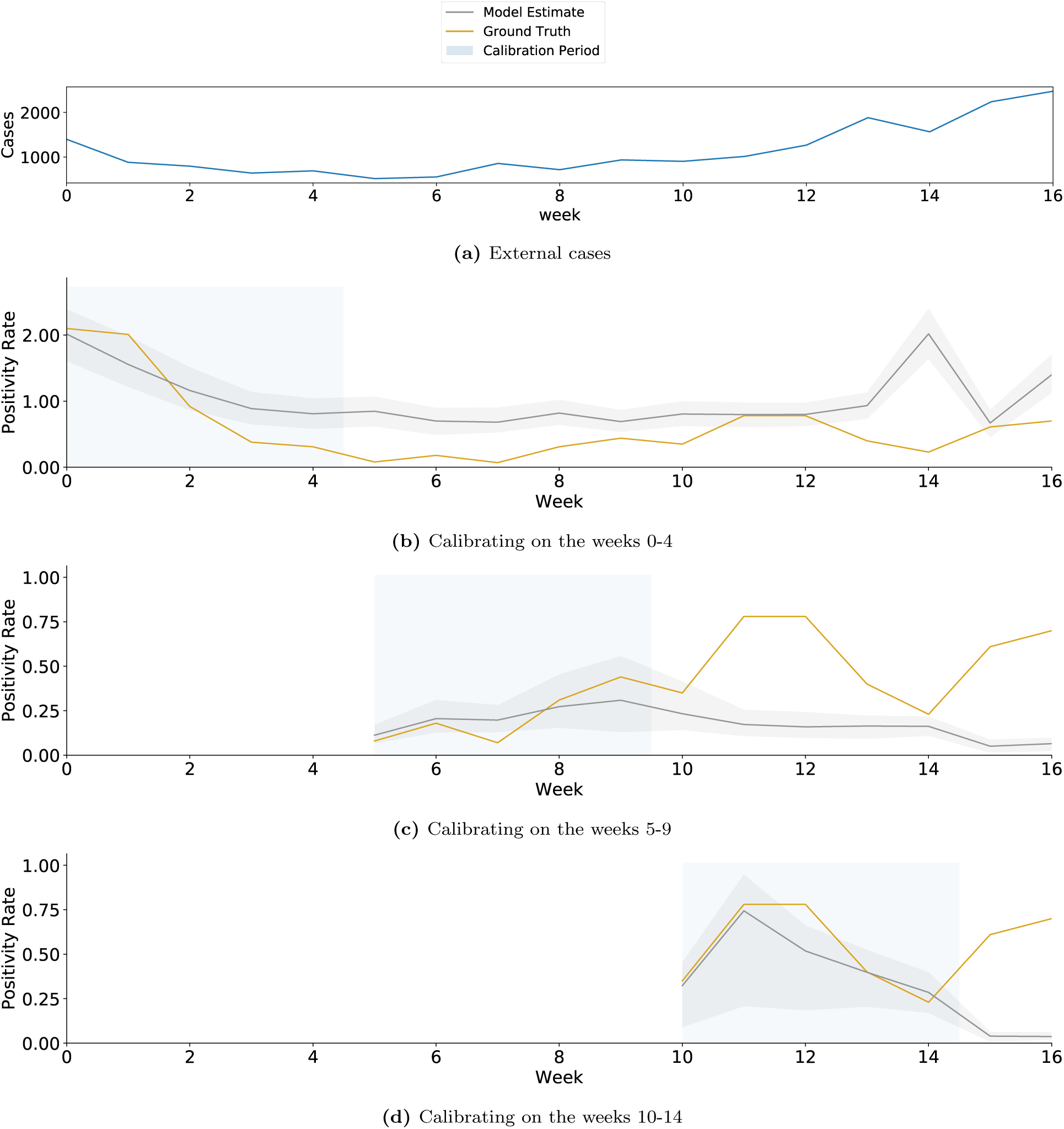
We calibrate ABM on positivity rates from Fall 2020 at GT. The objective function of the calibration is to minimize the r.m.s.e. with the weekly average of positivity rate obtained from surveillance testing results at GT [28]. (*a*) The parameter that determines external transmission of infections on a given day, *I_out_*(*t*), is a function of cases in Fulton county (where GT is located). (*b*) The models discussed in the main paper are calibrated using the first 5 weeks of data. We illustrate the output for a range of parameters that incorporate quantitative uncertainty, i.e., within 40% of the r.m.s.e. (*c, d*) illustrate calibration on the second period of 5 weeks and third period of 5 weeks respectively. These only show the optimal parameter output. The shaded region around the lines show the 2.5*^th^* and 97.5*^th^* percentile.

**Figure S4:**
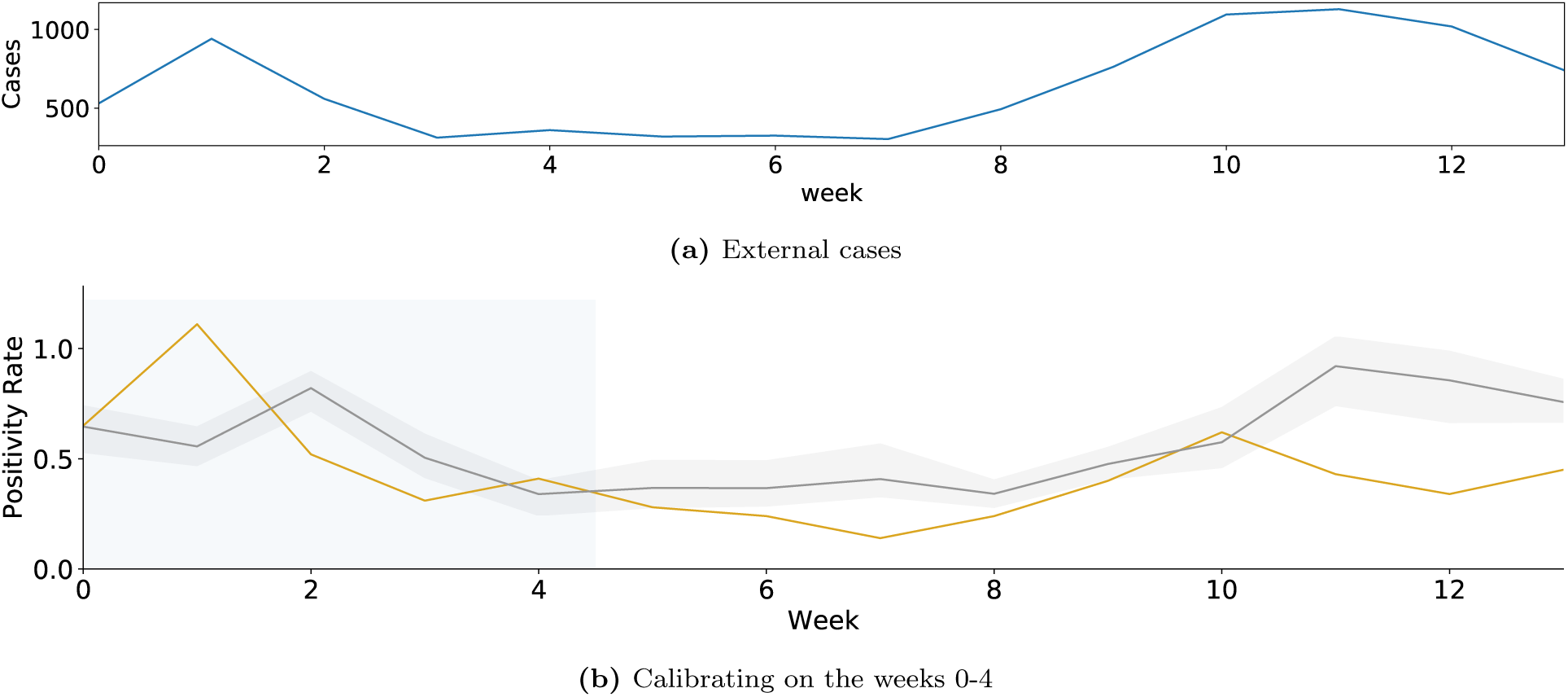
We calibrate ABM on positivity rates from first 5 weeks of Fall 2020 at UIUC. The objective function of the calibration is to minimize the r.m.s.e. with the weekly average of positivity rate obtained from surveillance testing results at GT [28]. (*a*) The parameter that determines external transmission of infections on a given day, *I_out_*(*t*), is a function of cases in Champaign county (where UIUC is located). (*b*) We illustrate the output for a range of parameters that incorporate quantitative uncertainty, i.e., within 40% of the r.m.s.e. The shaded region around the lines show the 2.5*^th^* and 97.5*^th^* percentile.

**Figure S6:**
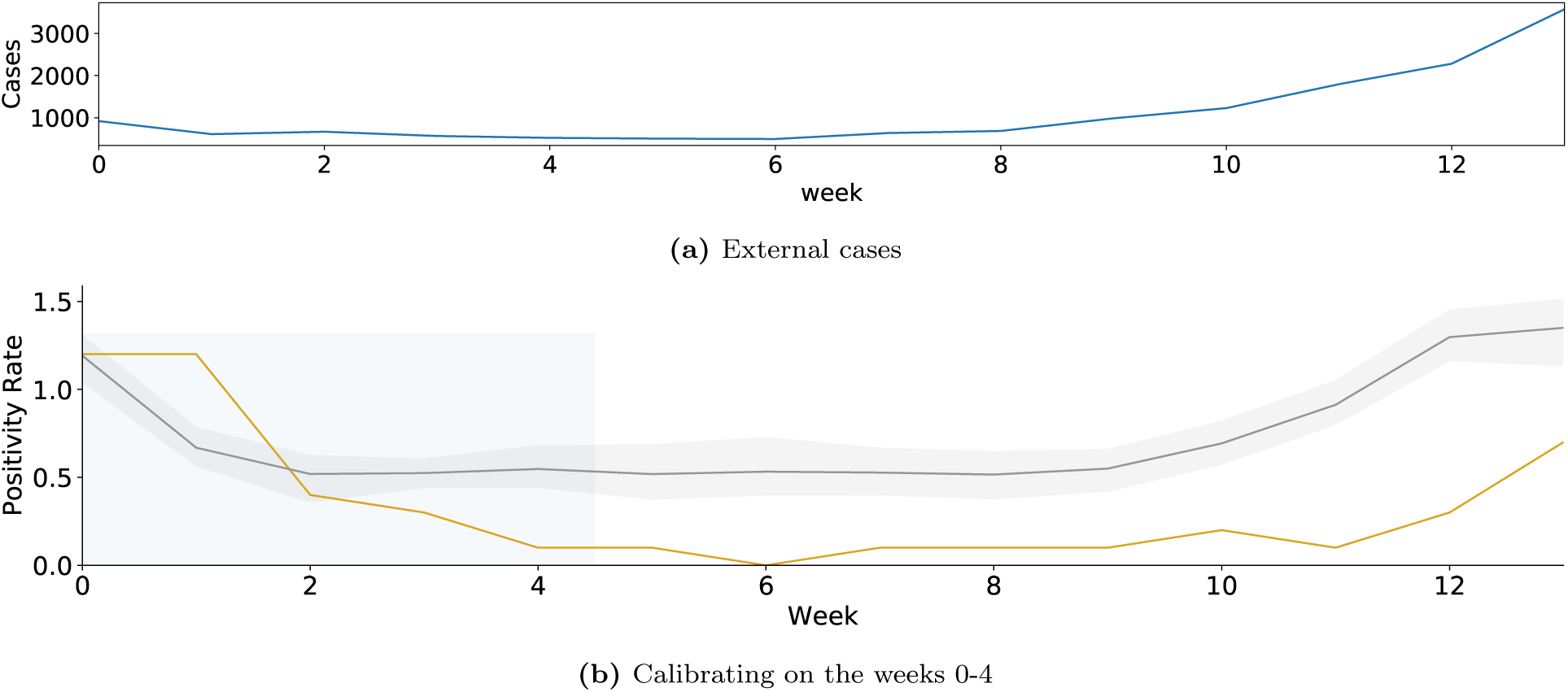
We calibrate ABM on positivity rates from first 5 weeks of Fall 2020 at UC Berkeley. The objective function of the calibration is to minimize the r.m.s.e. with the weekly average of positivity rate obtained from surveillance testing results at GT [28]. (*a*) The parameter that determines external transmission of infections on a given day, *I_out_*(*t*), is a function of cases in Alameda county (where UIUC is located). We illustrate the output for a range of parameters that incorporate quantitative uncertainty, i.e., within 40% of the r.m.s.e. The shaded region around the lines show the 2.5*^th^* and 97.5*^th^* percentile.

**Figure S7:**
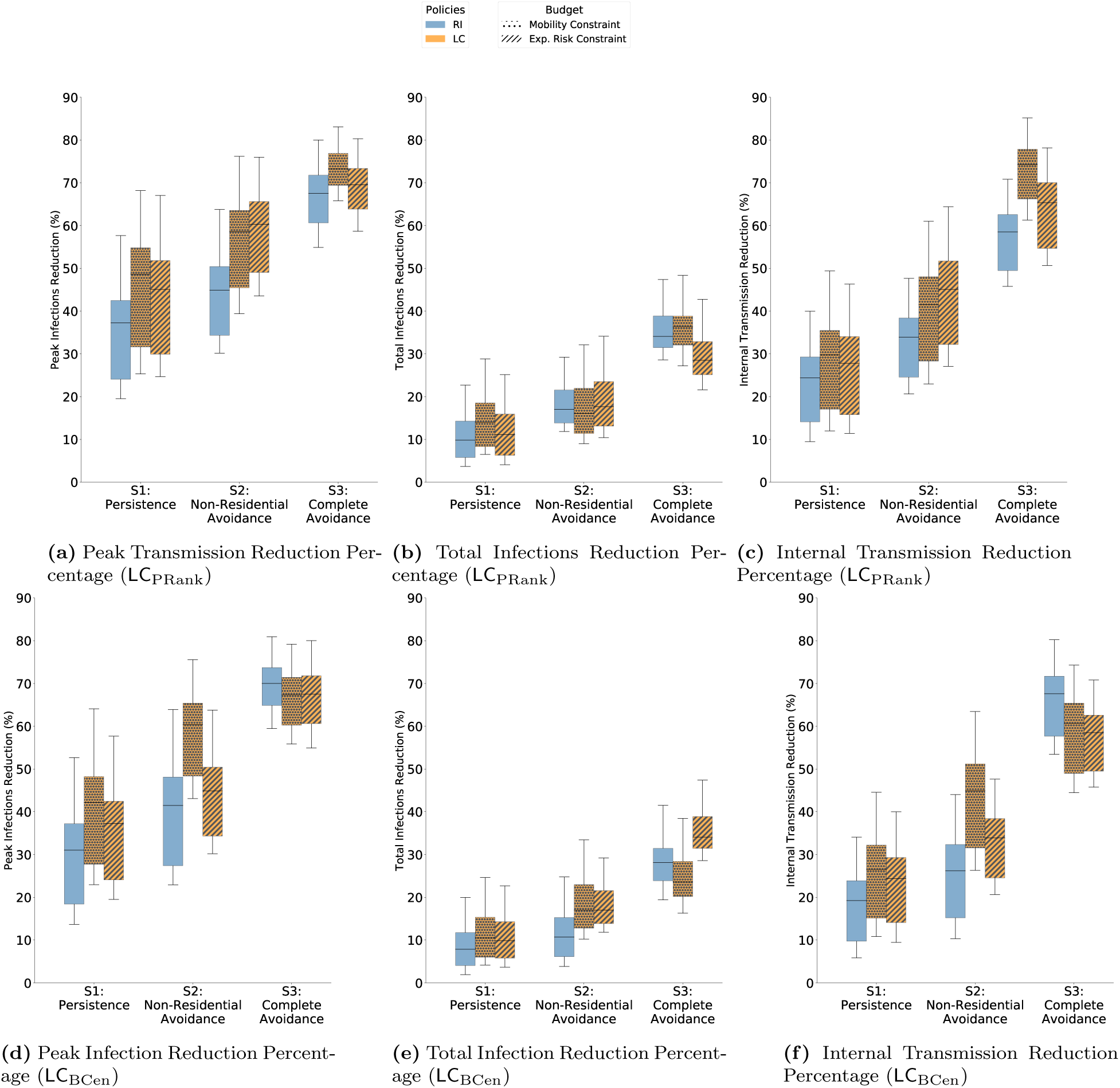
Disease control outcomes in Fall 2019 for different algorithms of LC with the ABM is calibrated on weeks 0−4 of Fall 2020 at GT. (*a − c*) Comparison of RI with LC_PRank_. Under all behavioral scenarios, for peak infection reduction (*b*) and internal transmission reduction (*c*), LC_PRank_ shows better disease control outcomes than RI. For total infection reduction (*b*), LC_PRank_ is better in *S1*, worse in *S3* when designed within an exposure risk budget, and comparable in others. (*d − f* ) Comparison of RI with LC_BCen_. Under all behavioral scenarios, for peak infection reduction (*d*) and internal transmission reduction (*f* ) LC_BCen_ is better when designed within an exposure risk budget. For total infection reduction (*e*), LC_BCen_ is always worse than RI

**Figure S8:**
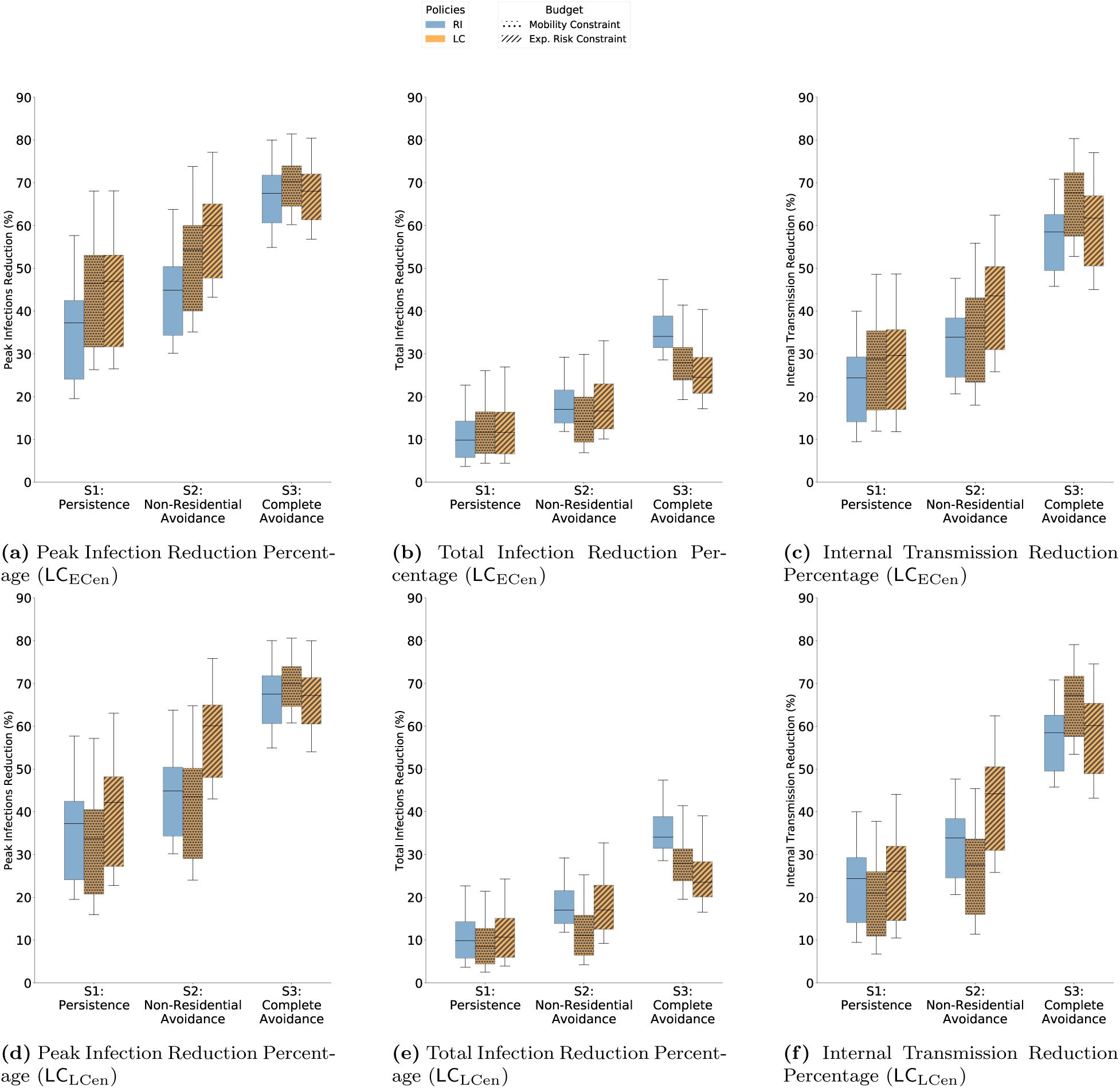
Disease control outcomes in Fall 2019 for different algorithms of LC with the ABM is calibrated on weeks 0 − 4 of Fall 2020 at GT. (*a − c*) Comparison of RI with LC_ECen_. Under all behavioral scenarios, for peak infection reduction (*b*) and internal transmission reduction (*c*), LC_ECen_ shows better disease control outcomes than RI. For total infection reduction (*b*), LC_ECen_ is better in *S1* and worse in *S3* when designed within an exposure risk budget. (*d − f* ) Comparison of RI with LC_ECen_. Under all behavioral scenarios, for peak infection reduction (*d*) and internal transmission reduction (*f* ), LC_ECen_ shows better disease control outcomes than RI. For total infection reduction (*e*), LC_ECen_ is better in *S1* and worse in *S3* when designed within an exposure risk budget.

**Figure S9:**
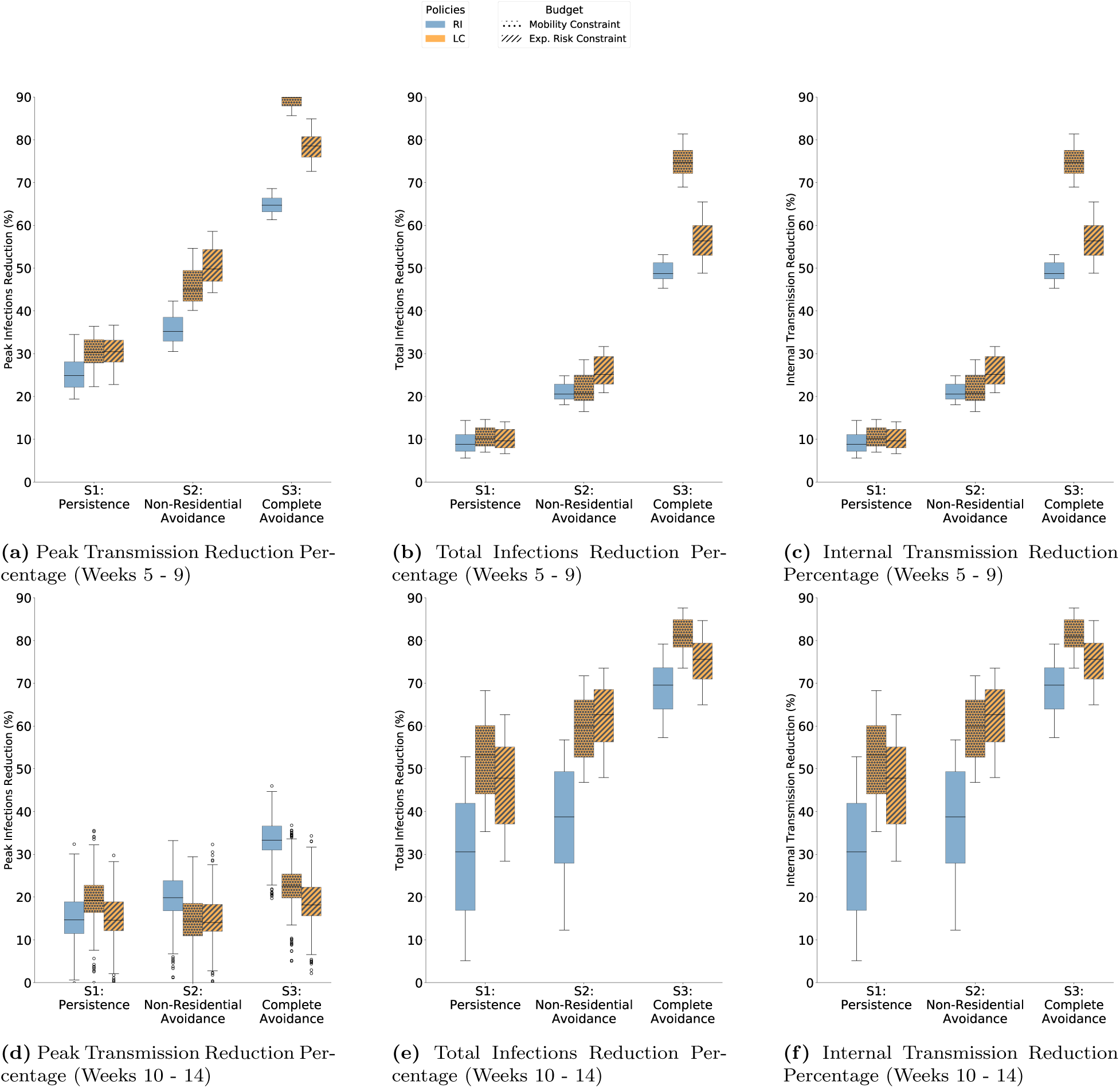
Disease control outcomes in Fall 2019 for LC_PRank_. (*a − c*) The ABM was calibrated on weeks 5 − 9 of Fall 2020 at GT. Under all behavioral scenarios, for all outcomes, LC_PRank_ is better than RI. (*d − f* ) The ABM was calibrated on weeks 10 − 14 of Fall 2020 at GT. Under all behavioral scenarios, for all outcomes, LC_PRank_ is better than RI.

**Figure S10:**
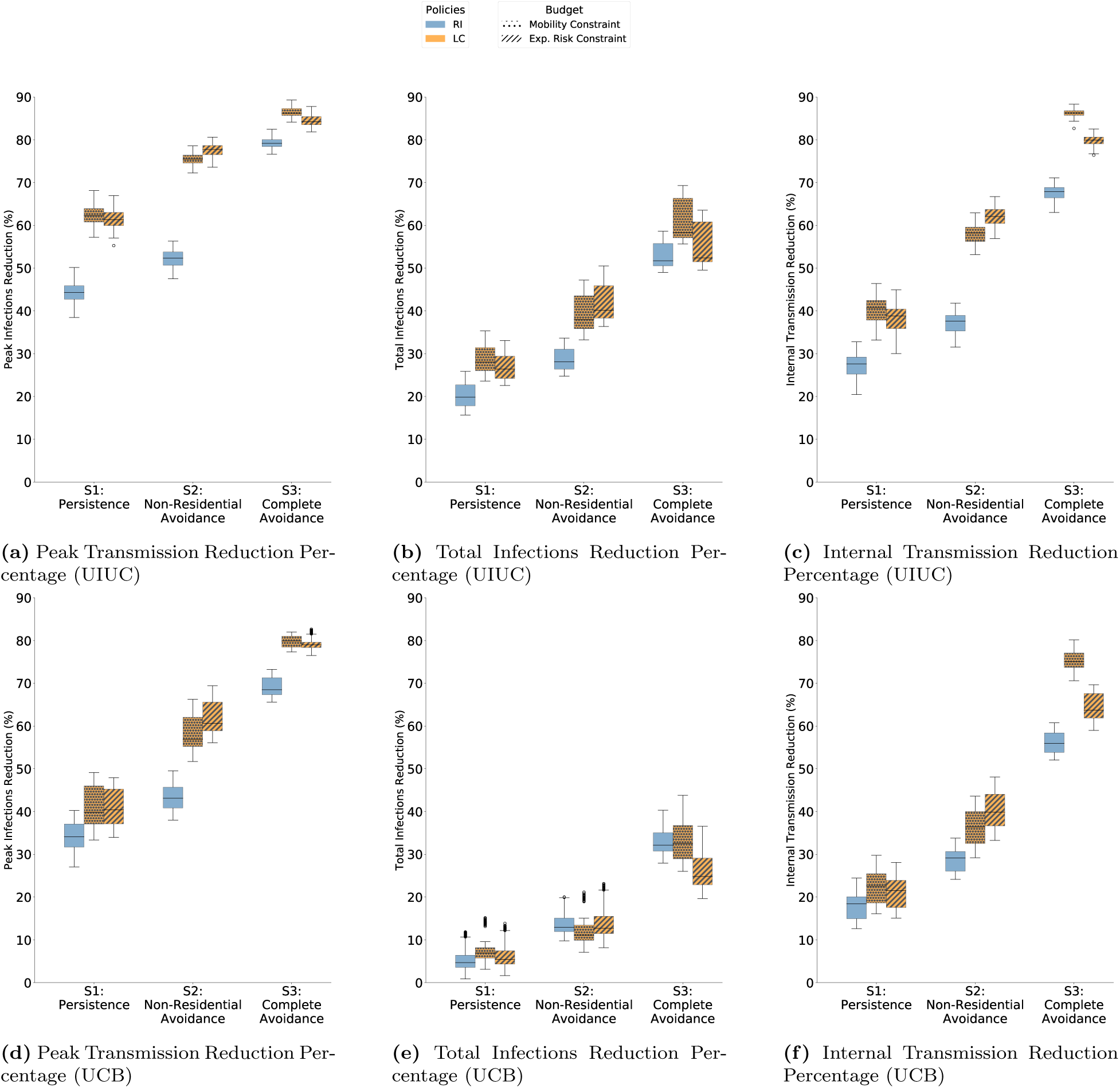
Disease control outcomes in Fall 2019 for LC_PRank_. (*a − c*) The ABM was calibrated on weeks 0 − 4 of Fall 2020 at UIUC. Under all behavioral scenarios, for all outcomes, LC_PRank_ is better than RI. (*d − f* ) The ABM was calibrated on weeks 0−4 of Fall 2020 at UC Berkeley. Under all behavioral scenarios, for all outcomes, LC_PRank_ is better than RI.

**Figure S11:**
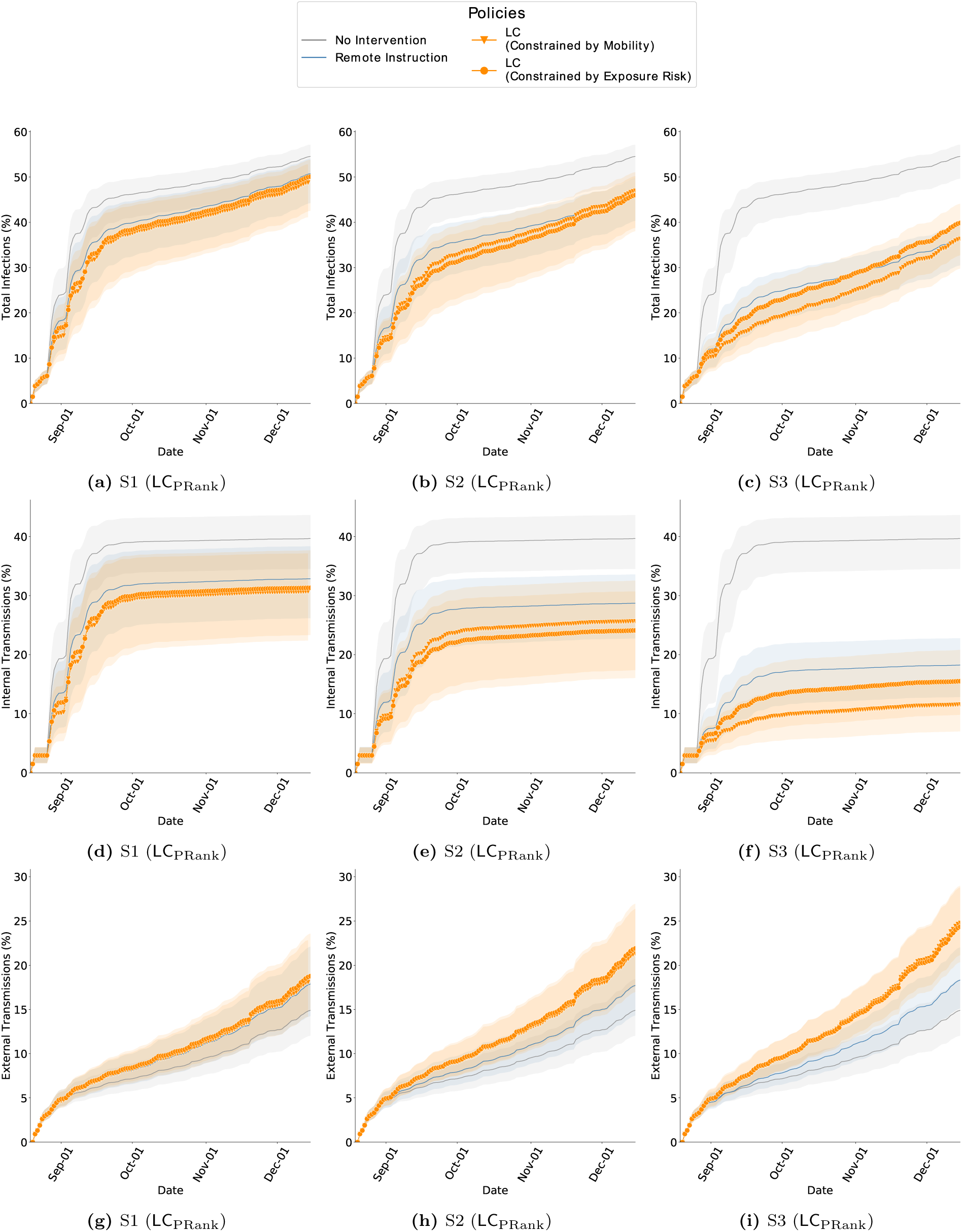
Cumulative infections in Fall 2019 while comparing RI and LC_PRank_ with ABM calibrated on weeks 0 *−* 4 of Fall 2020, GT. The bands show the 2.75*^th^* and 97.25*^th^* percentile. (*a − c*) Total infections of interventions is lower than no-intervention and is lowest in the *S3* scenario. In this behavioral scenario, the mobility budget is 69% of what it would be without interventions, and therefore the transmissions are also contained. In comparison, in Fall 2020, we saw far fewer infections which is because the mobility was 39% of that in Fall 2019. (*d − f* ) Internal transmissions are lower with LC_PRank_ in comparison to RI. (*g − i*) External transmissions are higher with LC_PRank_ in comparison to RI. Since internal transmission is controlled, more individuals remain susceptible to infections from outside campus.

**Figure S12:**
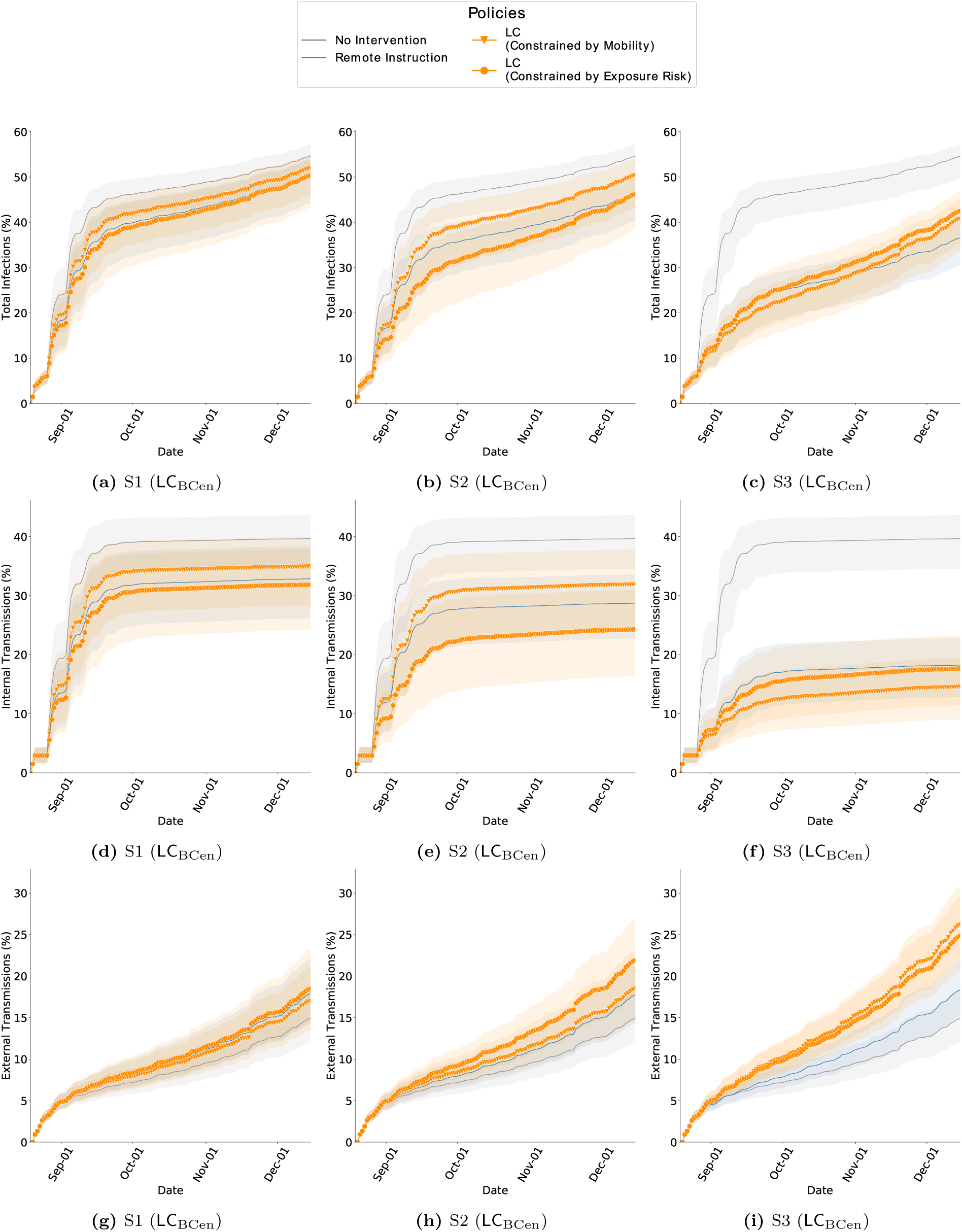
Cumulative infections in Fall 2019 while comparing RI and LC_BCen_ with ABM calibrated on weeks 0 *−* 4 of Fall 2020, GT. The bands show the 2.75*^th^* and 97.25*^th^* percentile. (*a − c*) Total infections of interventions is lower than no-intervention and is lowest in the *S3* scenario. In this scenario, the mobility budget is 69% of what it would be without interventions, and therefore the transmissions are also contained. In comparison, in Fall 2020, we saw far fewer infections which is because the mobility was 39% of that in Fall 2019. (*d − f* ) Internal transmissions are lower with LC_BCen_ in comparison to RI, only when constrained under the exposure risk budget. (*g − i*) External transmissions are higher with LC_BCen_ in comparison to RI. Since internal transmission is controlled, more individuals remain susceptible to infections from outside campus.

**Figure S13:**
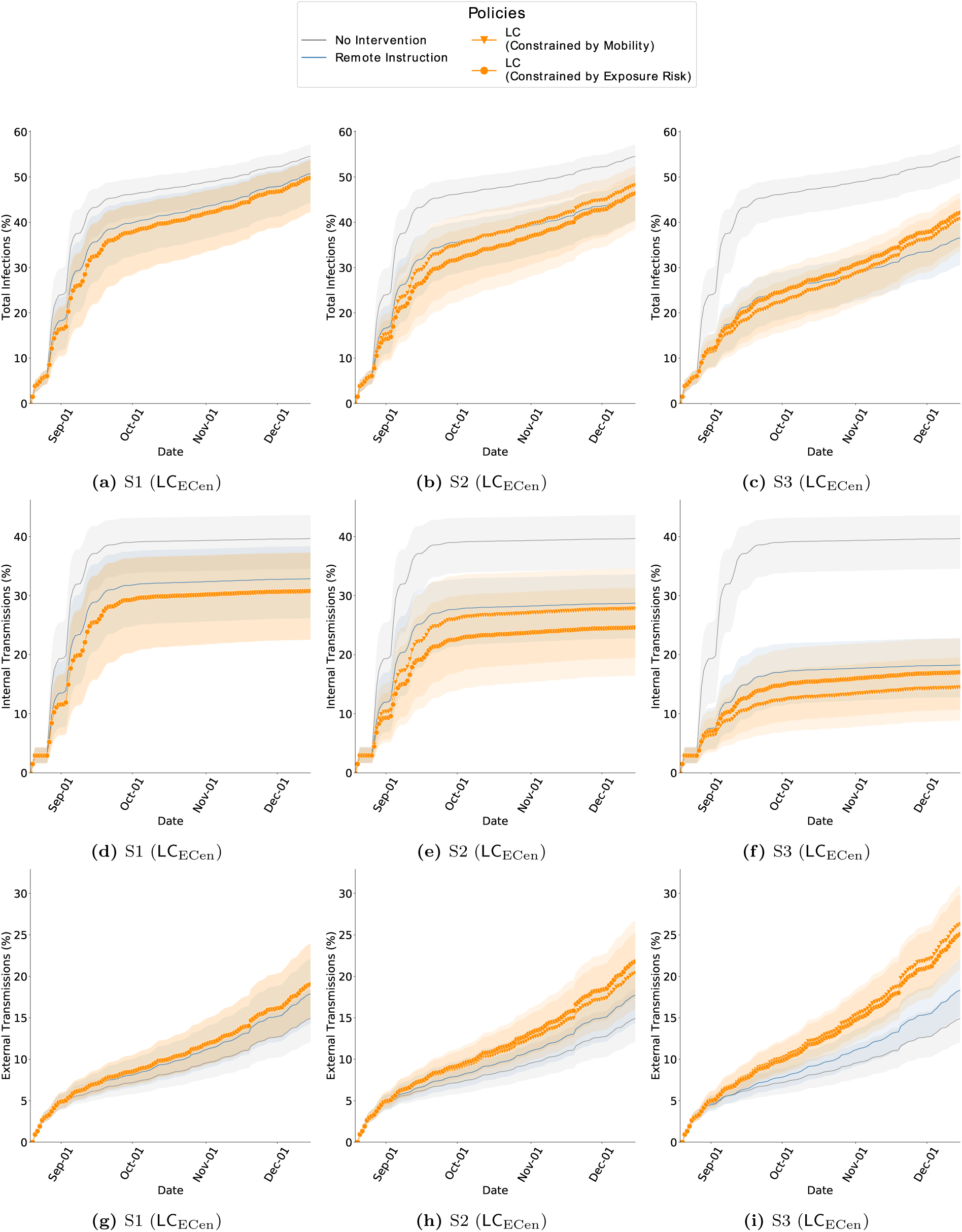
Cumulative infections in Fall 2019 while comparing RI and LC_ECen_ with ABM calibrated on weeks 0 *−* 4 of Fall 2020, GT. The bands show the 2.75*^th^* and 97.25*^th^* percentile. (*a − c*) Total infections of interventions is lower than no-intervention scenarios and is lowest in the *S3* scenario. In this scenario, the mobility budget is 69% of what it would be without interventions, and therefore the transmissions are also contained. In comparison, in Fall 2020, we saw far fewer infections which is because the mobility was 39% of that in Fall 2019. (*d − f* ) Internal transmissions are lower with LC_ECen_ in comparison to RI. (*g − i*) External transmissions are higher with LC_ECen_ in comparison to RI. Since internal transmission is controlled, more individuals remain susceptible to infections from outside campus.

**Figure S14:**
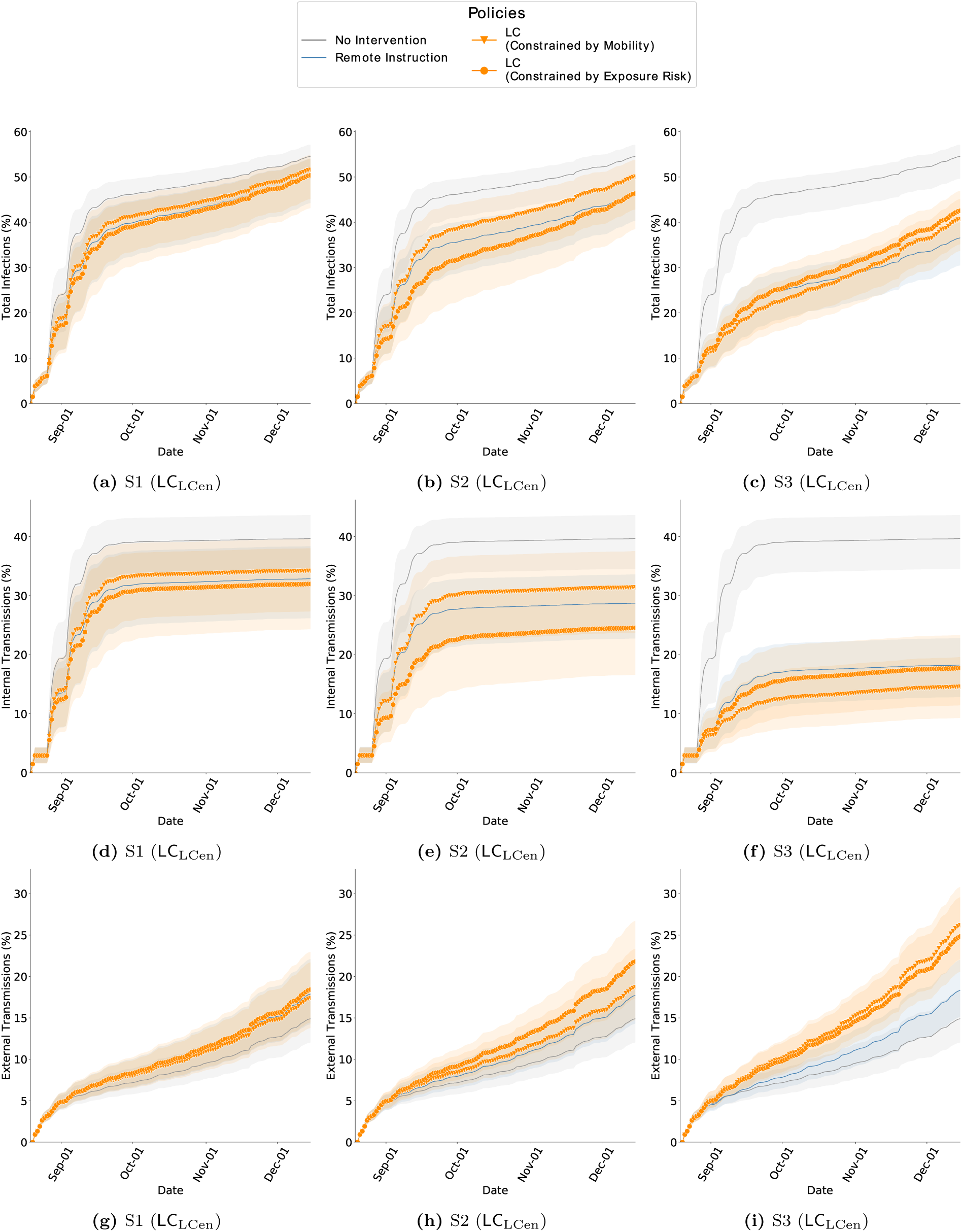
Cumulative infections in Fall 2019 while comparing RI and LC_LCen_ with ABM calibrated on weeks 0 *−* 4 of Fall 2020, GT. The bands show the 2.75*^th^* and 97.25*^th^* percentile. (*a − c*) Total infections of interventions is lower than no-intervention scenarios and is lowest in the *S3* scenario. In this scenario, the mobility budget is 69% of what it would be without interventions, and therefore the transmissions are also contained. In comparison, in Fall 2020, we saw far fewer infections which is because the mobility was 39% of that in Fall 2019. (*d − f* ) Internal transmissions are lower with LC_LCen_ in comparison to RI. (*g − i*) External transmissions are higher with LC_LCen_ in comparison to RI. Since internal transmission is controlled, more individuals remain susceptible to infections from outside campus.

**Figure S15:**
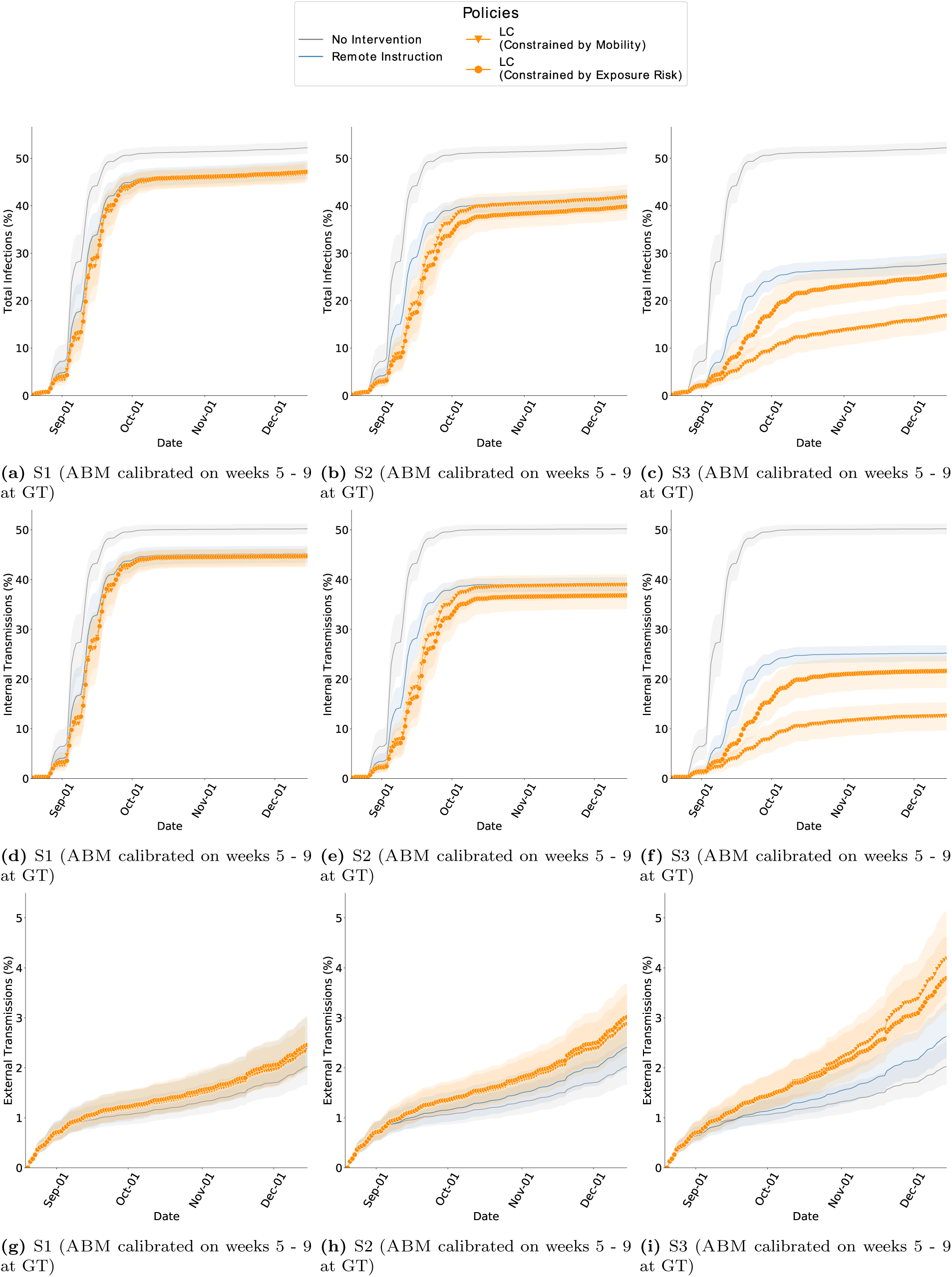
Cumulative infections in Fall 2019 while comparing RI and LC_PRank_ with ABM calibrated on weeks 5 *−* 9 of Fall 2020, GT. The bands show the 2.75*^th^* and 97.25*^th^* percentile. (*a − c*) Total infections of interventions is lower than no-intervention scenarios and is lowest in the *S3* scenario. In this scenario, the mobility budget is 69% of what it would be without interventions, and therefore the transmissions are also contained. In comparison, in Fall 2020, we saw far fewer infections which is because the mobility was 39% of that in Fall 2019. (*d − f* ) Internal transmissions are lower with LC_PRank_ in comparison to RI. (*g − i*) External transmissions are higher with LC_PRank_ in comparison to RI. Since internal transmission is controlled, more individuals remain susceptible to infections from outside campus.

**Figure S16:**
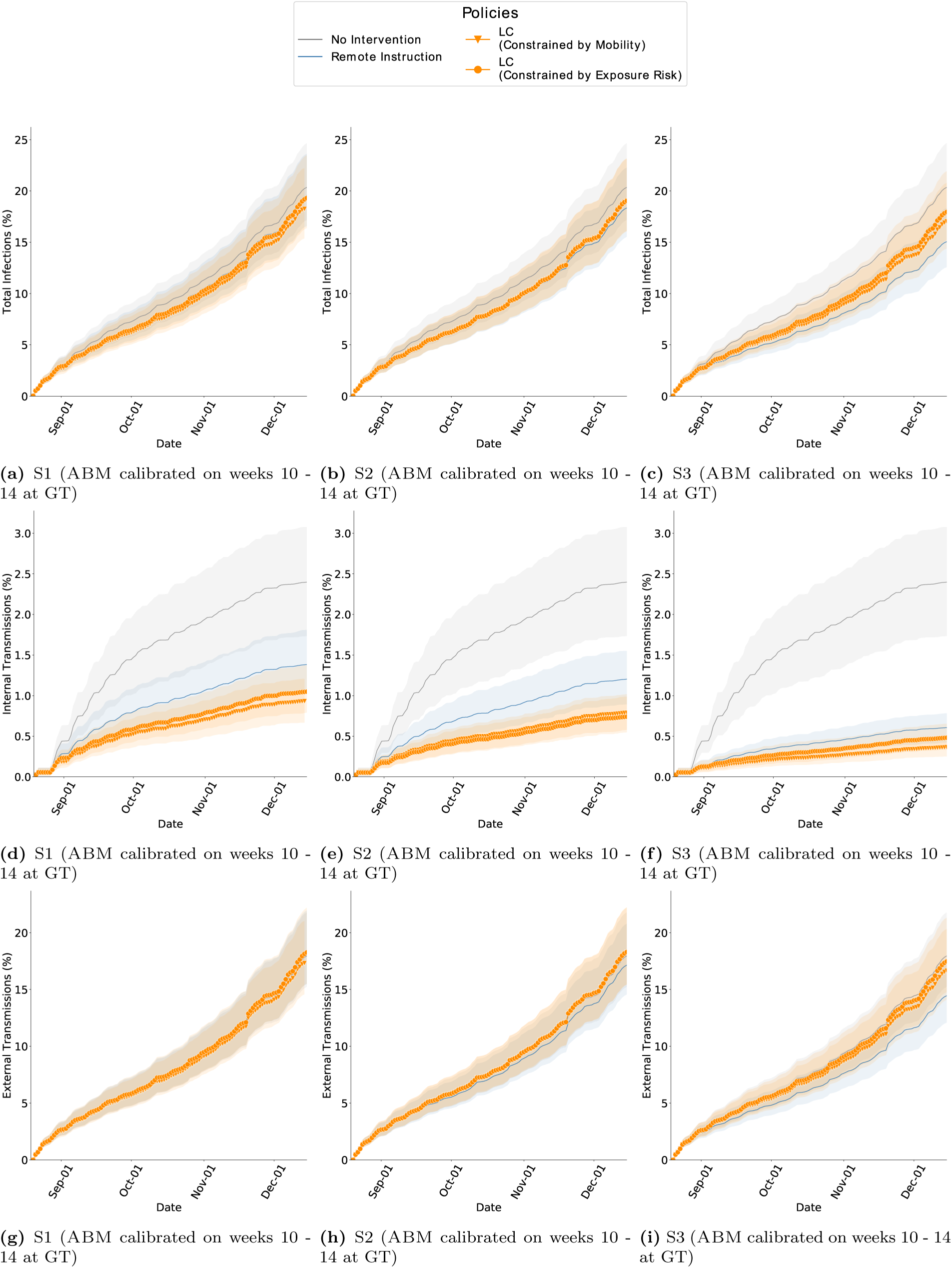
Cumulative infections in Fall 2019 while comparing RI and LC_PRank_ with ABM calibrated on weeks 10 *−* 14 of Fall 2020, GT. The bands show the 2.75*^th^* and 97.25*^th^* percentile. (*a − c*) Total infections of interventions is lower than no-intervention scenarios and is lowest in the *S3* scenario. In this scenario, the mobility budget is 69% of what it would be without interventions, and therefore the transmissions are also contained. In comparison, in Fall 2020, we saw far fewer infections which is because the mobility was 39% of that in Fall 2019. (*d − f* ) Internal transmissions are lower with LC_PRank_ in comparison to RI. (*g − i*) External transmissions are higher with LC_PRank_ in comparison to RI. Since internal transmission is controlled, more individuals remain susceptible to infections from outside campus.

**Figure S17:**
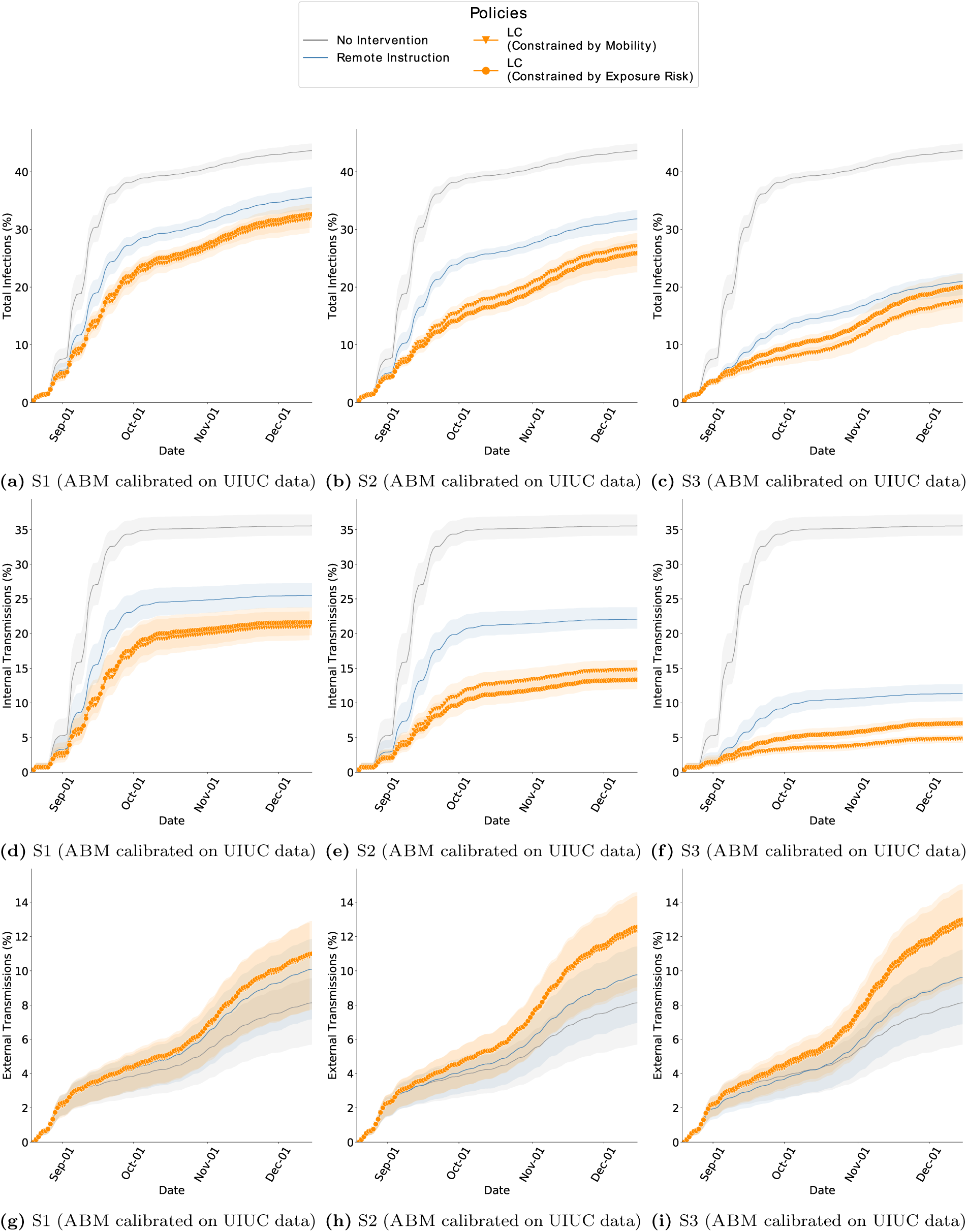
Cumulative infections in Fall 2019 while comparing RI and LC_PRank_ with ABM calibrated on weeks 0 *−* 4 of Fall 2020, UIUC. The bands show the 2.75*^th^* and 97.25*^th^* percentile. (*a − c*) Total infections of interventions is lower than no-intervention scenarios and is lowest in the *S3* scenario. In this scenario, the mobility budget is 69% of what it would be without interventions, and therefore the transmissions are also contained. In comparison, in Fall 2020, we saw far fewer infections which is because the mobility was 39% of that in Fall 2019. (*d − f* ) Internal transmissions are lower with LC_PRank_ in comparison to RI. (*g − i*) External transmissions are higher with LC_PRank_ in comparison to RI. Since internal transmission is controlled, more individuals remain susceptible to infections from outside campus.

**Figure S18:**
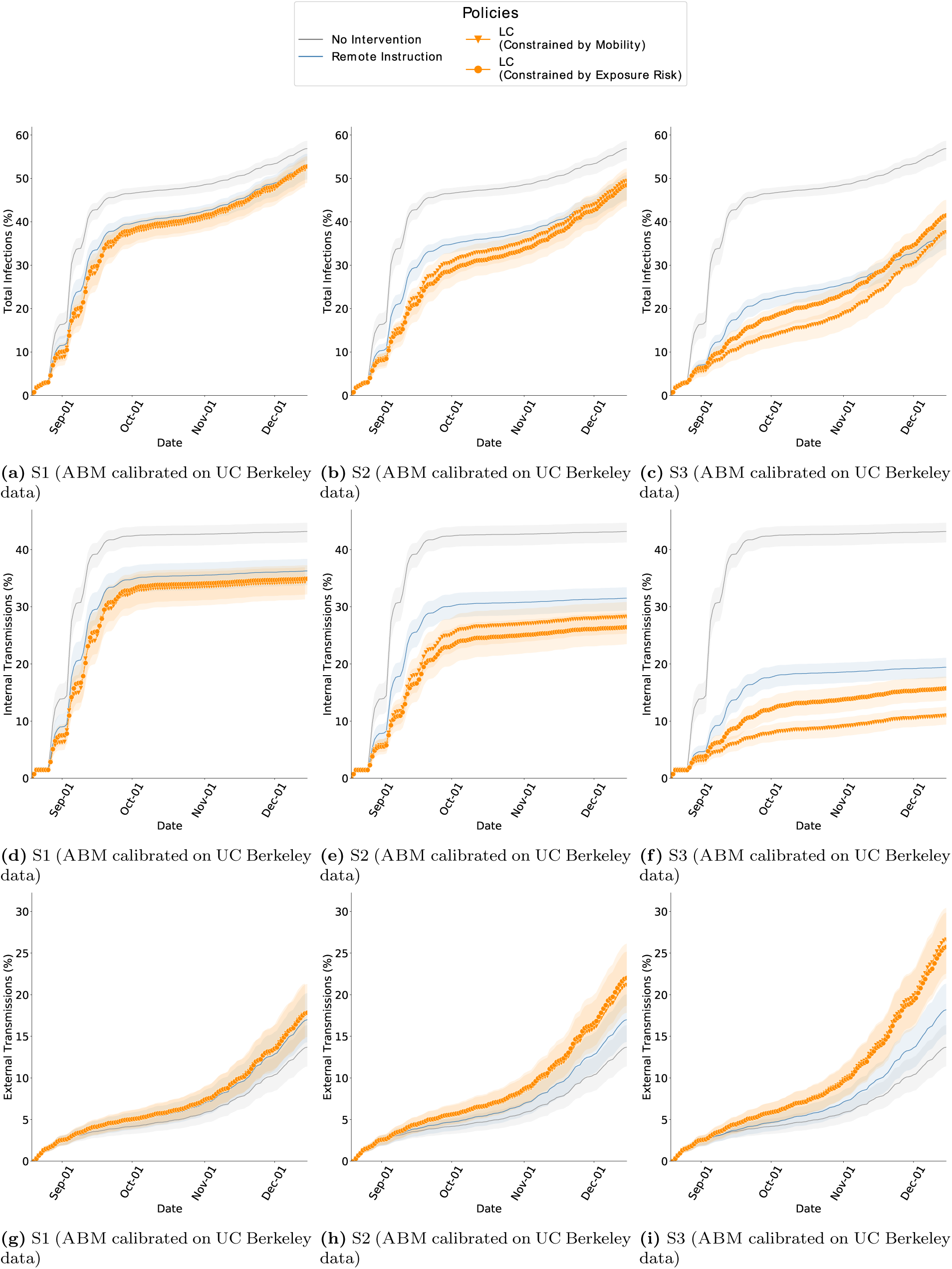
Cumulative infections in Fall 2019 while comparing RI and LC_PRank_ with ABM calibrated on weeks 0 *−* 4 of Fall 2020, UC Berkeley. The bands show the 2.75*^th^* and 97.25*^th^* percentile. (*a − c*) Total infections of interventions is lower than no-intervention scenarios and is lowest in the *S3* scenario. In this scenario, the mobility budget is 69% of what it would be without interventions, and therefore the transmissions are also contained. In comparison, in Fall 2020, we saw far fewer infections which is because the mobility was 39% of that in Fall 2019. (*d − f* ) Internal transmissions are lower with LC_PRank_ in comparison to RI. (*g − i*) External transmissions are higher with LC_PRank_ in comparison to RI. Since internal transmission is controlled, more individuals remain susceptible to infections from outside campus.

**Figure S19:**
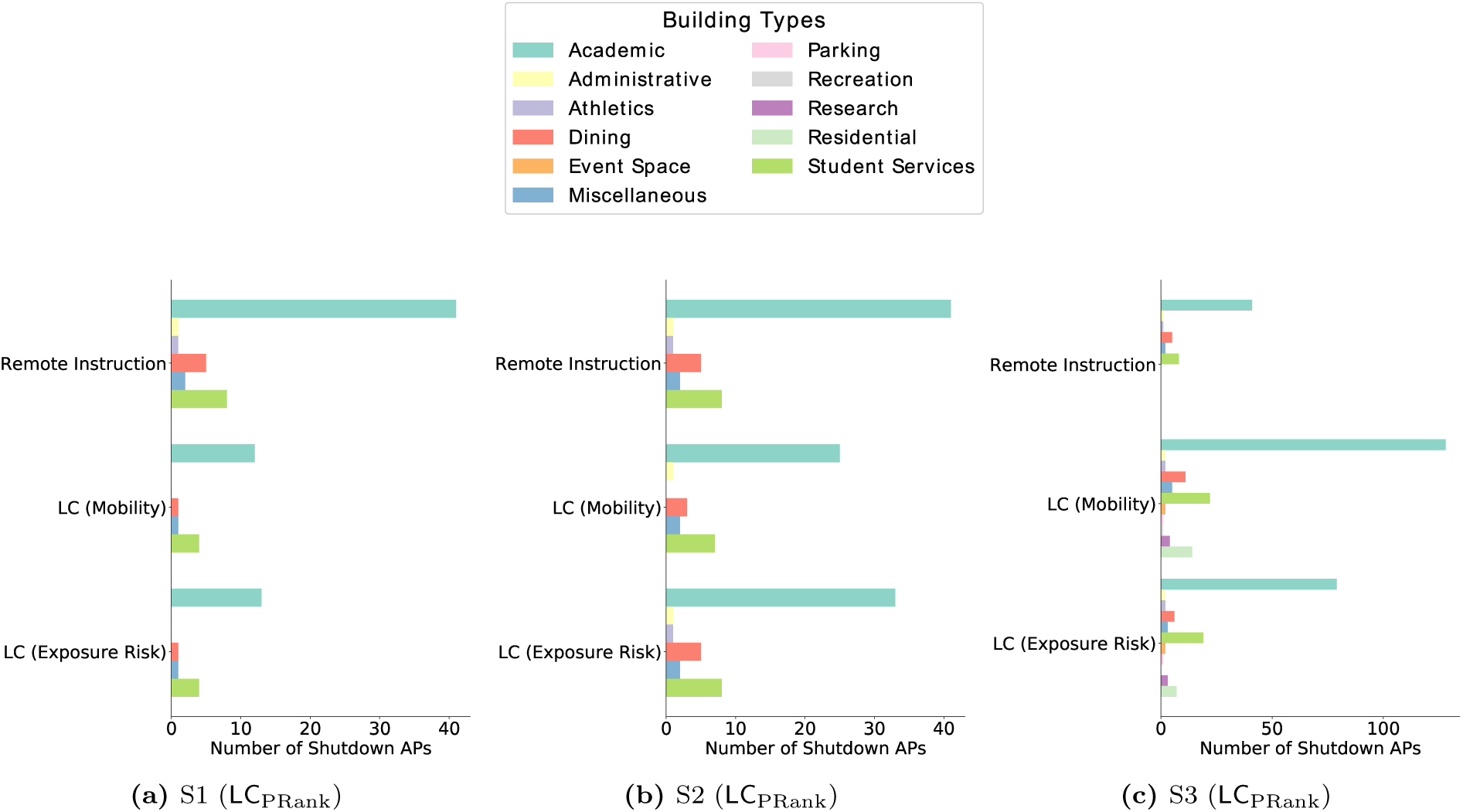
The locations shutdown by each policy are grouped into the the general building category. The distribution of locations is different between policies, for example, in *S1* (*a*) and *S2* (*b*), LC closes fewer locations that RI. Even when targeting spaces in similar buildings, the locations are qualitatively different RI only affects classrooms, whereas LC also closes smaller spaces like breakout rooms, reading areas and cafes. LC In *S3* (*c*) we find LC to target locations in a greater variety of buildings, but it also targets more locations to utilize the budget.

**Figure S20:**
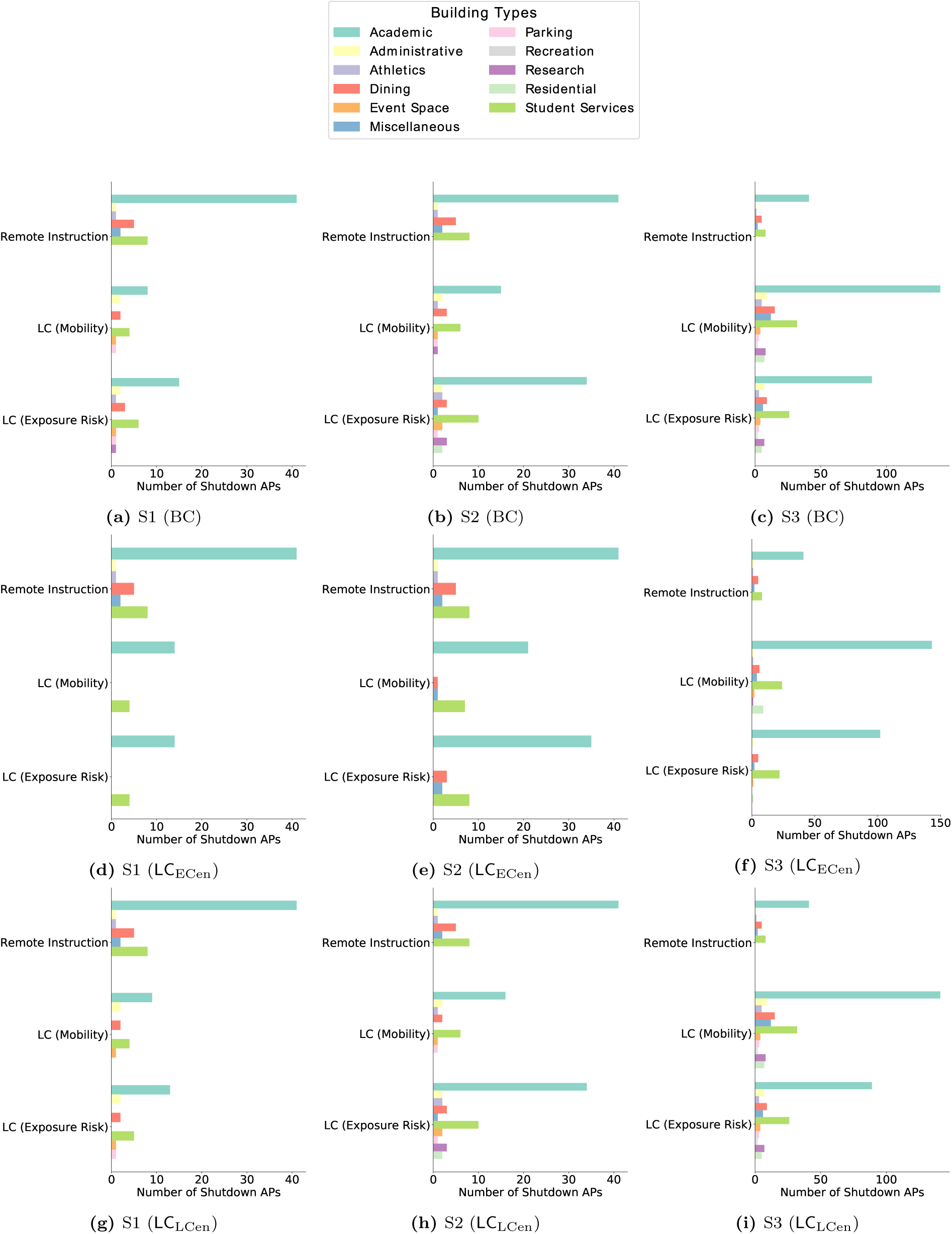
The locations shutdown by each policy are grouped into the the general building category. The distribution of locations is different between policies, for example, in *S1* (*a*) and *S2* (*b*), LC closes fewer locations that RI. Even when targeting spaces in similar buildings, the locations are qualitatively different RI only affects classrooms, whereas LC also closes smaller spaces like breakout rooms, reading areas and cafes. LC In *S3* (*c*) we find LC to target locations in a greater variety of buildings, but it also targets more locations to utilize the budget.

